# Epidemiologic shifts predict spatial patterns of measles re-emergence in the United States

**DOI:** 10.64898/2026.10.14.26365590

**Authors:** Chirag K. Kumar, Lauren A. Stoffel, Joshua L. Warren, Seth Blumberg, Robert Schechter, Kate M. Bubar, Mustafa Fattah, Mathew V. Kiang, Yvonne Maldonado, Erin A. Mordecai, Peter J. Hotez, Marc Lipsitch, Nathan C. Lo

**Affiliations:** Division of Infectious Diseases and Geographic Medicine, Department of Medicine, Stanford University, Stanford, CA, USA; Emmett Interdisciplinary Program in Environment and Resources, Doerr School of Sustainability, Stanford University, Stanford, CA, USA; Department of Biostatistics, Yale School of Public Health, New Haven, CT, USA; Department of Medicine and F.I. Proctor Foundation, University of California, San Francisco, San Francisco, CA, USA; California Department of Public Health, Richmond, CA, USA; Vagelos College of Physicians and Surgeons, Columbia University, New York, NY, USA; Department of Epidemiology and Population Health, Stanford University, Stanford, CA, USA; Departments of Pediatrics and Epidemiology and Population Health, Stanford University, Stanford, CA, USA; Department of Biology, Stanford University, Stanford, CA, USA; Texas Children’s Hospital Center for Vaccine Development, Department of Pediatrics and Molecular Virology and Microbiology, National School of Tropical Medicine, Baylor College of Medicine, Houston, TX, USA

**Keywords:** measles, vaccines, immunization, MMR, outbreak, epidemiology

## Abstract

Globally, measles is re-emerging in countries that have previously achieved elimination, including the United States. As immunization rates decline, characterizing the dynamics and evolving spatial pattern of measles re-emergence is critical to inform the public health response. We analyze annual county-level measles cases and vaccine coverage data in the United States (US) from 2011-2025 and find a shift in measles epidemiology in recent years indicative of re-emergence. While outbreak risk is spatially concentrated, we find that outbreaks are increasingly spreading throughout the broader population and across counties, but targeted interventions are predicted to prevent re-emergence nationwide. During the first decade of this period (2011-2020), outbreaks were only weakly associated with county-level vaccine coverage and predominantly isolated to single counties; however, recently (2021-2025), vaccine coverage has become strongly associated with outbreak size and spread across neighboring counties has increased. These findings suggest that measles transmission is increasingly sustained across the broader population rather than only in socially clustered communities. We developed a spatial statistical model to predict outbreak risk and size in all US counties with validation over 2021-2025; the model predicted counties with ≥10 cases with a sensitivity of 95% (range across 2021-2025: 75-100%) and specificity of 83% (64-98%). Mechanistic models based on county vaccine coverage alone had worse performance, particularly in predicting larger outbreaks. Using the spatial statistical model, we find that prioritizing interventions in the areas of greatest model-predicted risk (e.g., top decile of US counties) was more impactful than state-level interventions (e.g., top five states) or targeting based on low vaccine coverage alone. Achieving even modest (5 percentage point) increases in vaccine coverage in counties in the top decile of model-predicted risk reduced long-term measles risk nationwide by over 90%. Together, these findings demonstrate that predictive risk modeling to inform spatially targeted interventions can likely mitigate the re-emergence of vaccine-preventable diseases.

## Main Text

In recent years, measles has re-emerged around the world in settings where the disease had been previously eliminated,^1–4^ resulting in multiple countries losing their World Health Organization measles elimination status.^3^ Global measles incidence has risen sharply,^4^ driven by growing vaccine hesitancy^5–7^ and disruptions to immunization programs during the COVID-19 pandemic.^8^ In the United States (US), the disease was declared eliminated in 2000, but there have recently been a series of large and sustained outbreaks that threaten this elimination status.^2,9,10^ Because measles is among the most transmissible human pathogens, even small gaps in immunity can lead to significant outbreaks,^11–15^ but factors beyond vaccine coverage alone also impact where outbreaks occur and their size.^16,17^ While cases occur largely among unvaccinated individuals,^9^ measles re-emergence risk is a complex interplay between immunity, exposure to travel-acquired internationally imported cases, population structure,^18^ and the public health response.^19^ Quantifying the interplay of these factors as measles re-emerges in elimination settings is critical to characterize the dynamics and evolving spatial distribution of re-emergence risk to inform prioritization of interventions in the most susceptible locations.

There are limited data on the epidemiologic patterns of measles re-emergence in post-elimination settings.^20^ While the dynamics behind endemic measles^21–25^ and the drivers of eliminating transmission across settings^26–30^ have been well characterized, understanding the dynamics of re-emergence remains challenging. Following measles elimination in the US, outbreak risk has often been assumed to be a function of vaccine coverage alone,^31–33^ but vaccine coverage even at the county-level can obscure shifts in susceptibility that drives outbreak risk.^34^ Post-elimination measles outbreaks have been predominantly concentrated among small, socially clustered communities that are challenging to identify.^35^ However, as measles outbreaks have increased in the US, characterizing the relationship between county-level indicators and outbreak size is important for evaluating whether transmission is expanding beyond just tightly knit communities and instead predicted by the broader population’s vaccine coverage, indicative of an epidemiologic shift towards re-emergence.

Evaluating what models best predict outbreak risk could allow targeted, pre-emptive public health measures, including supplemental vaccination efforts, community-based outreach to under-immunized groups, or policy responses related to childhood vaccine requirements.^36^ To increase vaccine coverage, some US states have passed laws to remove nonmedical (i.e., religious or personal belief) exemptions to childhood vaccines^37–39^ and local health departments have engaged in targeted outreach and vaccination campaigns in areas with low coverage.^40^ However, predicting outbreak risk remains challenging,^41^ requiring estimation of both internationally imported cases due to travel^42^ and onwards local transmission risk,^43^ and public health response has therefore remained largely reactive.

Here, we study the epidemiologic patterns associated with re-emergent measles outbreaks in the US and develop spatial risk models to identify counties at high risk for outbreaks amid declining vaccine coverage. We characterize the shifting epidemiology of measles transmission in the US and synthesize these insights in validated predictive models that project county-level measles cases under plausible future vaccine coverage scenarios and policy interventions. Our findings can inform public health response to mitigate the risk of re-emergence of vaccine-eliminated diseases as childhood immunization rates decline.

## Results

### Epidemiologic shift associated with measles re-emergence in the United States

We assembled a US county-level measles case dataset for all county-years from 2011-2025, obtaining county-level assignment for >95% of CDC-reported state-level cases for a total of 5,739 cases with ≥1 cases in 848 county-years (Figure 1a, 1b; Figure S1; Data missingness is described in the Appendix). 10.0% of county-years had ≥10 cases, and cases in these counties accounted for 73.8% of total cases. We obtained county-level MMR vaccine coverage (or alternatively, up-to-date vaccine coverage where the child had to receive all vaccines and doses on their state’s list to meet criteria for being vaccinated; see Appendix) among entering schoolchildren (mainly kindergarteners), nonmedical exemption prevalence, and medical exemption prevalence for the same time period.^44,45^ MMR vaccine coverage among entering schoolchildren declined from a county average of 92.9% in 2011-2016 to 91.3% in 2025. In 2025, 67.4% of US counties with vaccine coverage data were below the critical threshold of 95% coverage (Figure 1c), and county-level vaccine coverage declined at an average rate of 0.38 percentage points annually from 2021-2025 (Figure 1d). We had vaccine data for 73.3% of county-years with measles cases (80.5% of cases), which was the main dataset for inferential analyses. We performed sensitivity analyses with imputed county-level vaccine data from adjacent years or state-level data from that year, increasing available vaccine data to 91.4% of county-years with measles cases (95.4% of cases).

**Figure 1:**
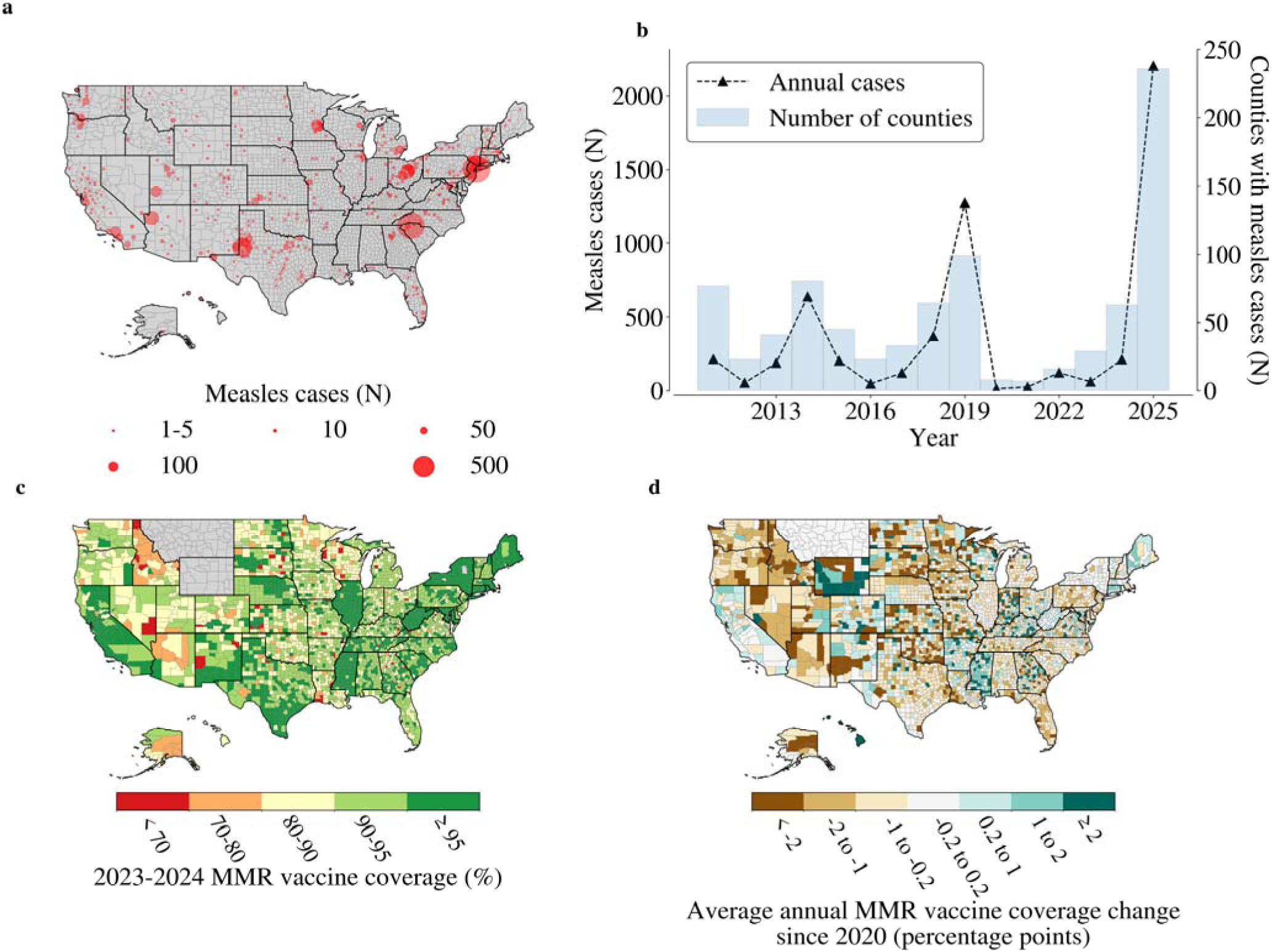
Reported measles cases and MMR vaccine coverage and trends in the US. (a) Map of county-level reported measles cases (internationally imported and locally acquired) used in this analysis, 2011-2025. The area of the red point is proportional to the total number of measles cases reported in the county over the entire study period. (b) Measles cases over time. Black triangles show reported measles cases by year (left side y-axis), and the blue histogram bars show the number of counties reporting at least one case in that year (right side y-axis). (c) Map of reported MMR vaccine coverage in school-aged children (mostly entering kindergartners) for 2023-2024 by county. Grey counties did not report data in 2023-2024. (d) Map of average rate of change in MMR vaccine coverage from 2020 by county. Grey counties did not report at least two years of data in the 2020-2025 time period.

We found that the relationship between county-level MMR vaccine coverage and measles outbreaks has changed over time. Using a hierarchical Bayesian Poisson model, we analyzed the relationship between county-level vaccine coverage and outbreak size (the number of reported annual measles cases minus known internationally imported cases) after introduction of a measles cases, adjusting for state– and year-variability, sociodemographic covariates (such as population density, income, demographics, and education; see Methods for full list of covariates), and spatial relationships between counties. We allowed the relationship between outbreak size and vaccine coverage to vary by year and found a significant nationwide strengthening in this relationship over time (Spearman ρ = – 0.70 (95% CrI: –0.91, –0.35); *p =* 0.003) with vaccine coverage becoming a significant negative predictor (i.e., lower county-level vaccination associated with larger outbreak sizes) in 2021 (Figure 2a), indicating a shift in the epidemiology of measles outbreaks in recent years. These findings were robust to various alternate model parameterizations of the time-varying relationship between vaccine coverage and outbreak size (Figure S2), supporting the hypothesis that recent outbreaks reflect transmission more broadly across the general population as quantified through county-level vaccine coverage rather than clusters of underimmunized individuals obscured by county-level coverage alone.

**Figure 2:**
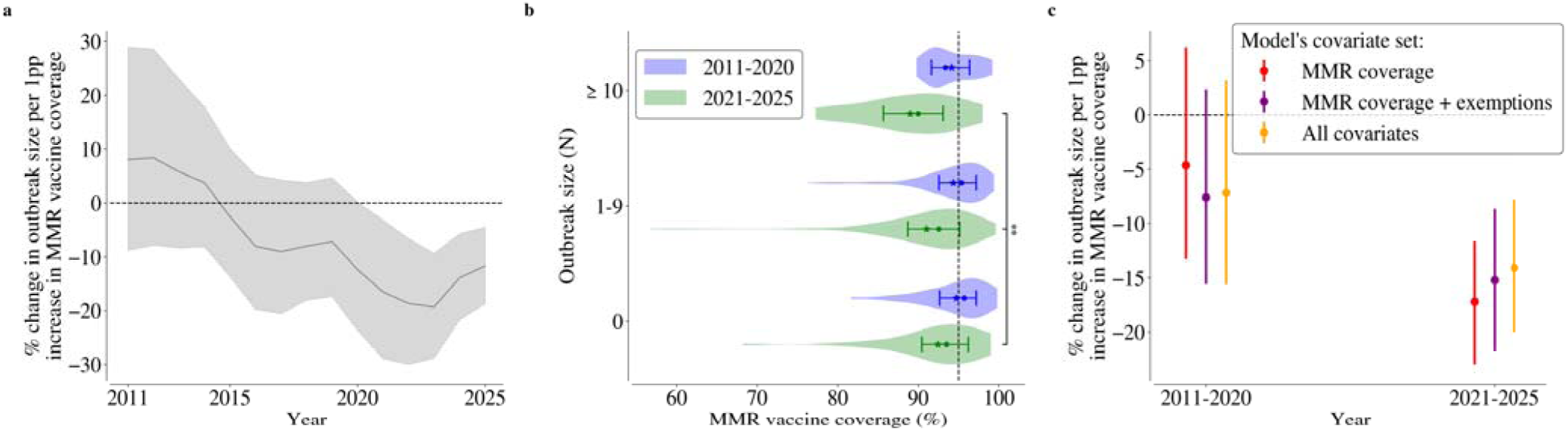
Relationship between county-level MMR vaccine coverage and measles outbreaks has changed over time. (a) Changing relationship between MMR vaccine coverage and measles outbreak size over time. We used Poisson models that adjusted for state– and year-variability as well as sociodemographic covariates and a conditional autoregressive spatial random effect at the county-level to allow the relationship between MMR vaccine coverage, exemptions, and demographic control covariates to vary over time. The line shows the posterior median of the estimated nationwide relationship between vaccine coverage and outbreak size over time with outliers removed (Figure S2e), and the shaded area shows the 95% credible interval. The measles outbreak size is quantified as the number of reported annual measles cases minus number of confirmed internationally imported cases. The curve demonstrates a strengthening relationship between vaccine coverage and outbreak size becoming significant in 2021. (b) County-level MMR vaccine coverage by measles outbreak size from 2011-2020 compared to 2021-2025 among county-years with at least one measles case introduction. An outbreak size of zero indicates only imported cases without secondary cases. Data were reported as annual county-level case counts, without confirmation that cases were epidemiologically linked. Points show the median MMR vaccine coverage across observations in each category, stars show the mean, and error bars show the 25^th^ and 75^th^ percentiles. The vertical dashed line is drawn at 95% vaccine coverage, the targeted critical immunity threshold in the US. The asterisks indicate that vaccine coverage and outbreak size were significantly correlated in the 2021-2025 period (Spearman ρ = –0.17; *p* = 0.009) but not in the 2011-2020 period. (c) Estimate for the impact of county-level MMR vaccine coverage on measles outbreak size using Poisson regressions with different sets of covariates (colors) fit separately to outbreak data from 2011-2020 and 2021-2025. All models include state– and year-fixed effects and a conditional autoregressive spatial random effect at the county level. Coefficients for MMR coverage, nonmedical exemptions, and medical exemptions can vary by state to account for state-level variation (Figure S1) and reporting differences. Points show the posterior median of the estimated nationwide relationship between MMR vaccine coverage and outbreak size, and error bars show the 95% credible interval.

We replicated the finding of a strengthening relationship between vaccine coverage and outbreak size over time when separately analyzing outbreaks across two time periods of 2011-2020 and 2021-2025, dividing the data based on the epidemiologic shift observed in 2021 when the vaccine coverage-outbreak size relationship becomes significant (Figure 2a). Prior to 2021, after introduction of a measles cases, county-level vaccine coverage and outbreak size (where zero indicates that there was no transmission from any imported cases) were not correlated (Spearman ρ = –0.09; *p* = 0.2) (Figure 2b). Since 2021, outbreak size after introduction of a measles case was significantly and more strongly correlated with vaccine coverage (Spearman ρ = –0.17; *p =* 0.009), underscoring that the epidemiological indicators of measles outbreaks have changed as re-emergence has occurred (Figure 2b; Figure S3-4). Accordingly, we estimated that a one percentage point increase in MMR vaccine coverage corresponded to a 14.1% (7.8, 20.0) decrease in outbreak size or a 1.3 (0.7, 1.9) reduction in cases for the average outbreak from 2021-2025 (Figure 2c).

The strengthening relationship between vaccine coverage and outbreak size is robust under various sensitivity analyses. The impact of vaccine coverage on outbreak size was consistent across states and vaccine coverage (Figure S5), indicating a nationwide-consistent shift in the epidemiology of measles outbreaks. Similarly, we replicated this relationship even after accounting for whether there were prior outbreaks in the county (Figure S6) and when using imputed vaccine coverage from neighboring years or state-level data in counties with measles cases but where vaccine data for that ear was unavailable (Figure S7). Finally, instead of using the vaccine coverage in the outbreak year, we used the average vaccine coverage over the preceding five years in a county, reflecting immunity over a larger childhood age group but diminishing the impact of recent changes in vaccine coverage, and found that this measure showed reduced sensitivity to the strengthening relationship between vaccine coverage and outbreak size, emphasizing the influence of recent declines in vaccine coverage on this shift (Figure S8).

Despite the increasing association between vaccine coverage and outbreak size, explaining outbreak size required additional sociodemographic factors and incorporating spatial relationships between counties. We calibrated our hierarchical Bayesian Poisson regression model to annual measles cases in counties with at least one case (i.e., an importation) from 2021-2025 and compared it against a benchmark mechanistic model that estimated outbreak size using final outbreak size calculations from vaccine coverage, enabling a mechanistically-grounded testing of the extent to which vaccine coverage alone predicts outbreak risk after controlling for state– and year-fixed effects. The hierarchical Bayesian Poisson regression model had better calibration performance than the mechanistic model (Widely-Applicable Information Criterion of 1360.2 vs 7857.8; Figure 3a; validation performance presented in Table 1 and Figure 4b). Both models had better calibration performance in 2021-2025 than 2011-2020 (Figure S9), reflecting that outbreaks have become increasingly characterizable. The associations between the county-level sociodemographic variables in the hierarchical Bayesian Poisson regression and outbreak size have also changed over time (Figure 3b). Income switched directionality between the two time periods: from 2011-2020, higher incomes were associated with larger outbreaks while during 2021-2025, lower incomes were associated with larger outbreaks. The median association between additional covariates, such as education levels, and outbreak size also switched directionality between the two time periods. Outbreak size was more strongly associated with the fraction of the population that is school aged in 2021-2025 than 2011-2020, and there is a consistent lower risk association with public schooling in both time periods (Figure S9d). Nonmedical exemptions were an insignificant predictor from 2021-2025 likely because of their association with vaccine coverage (Figure S10).

**Figure 3:**
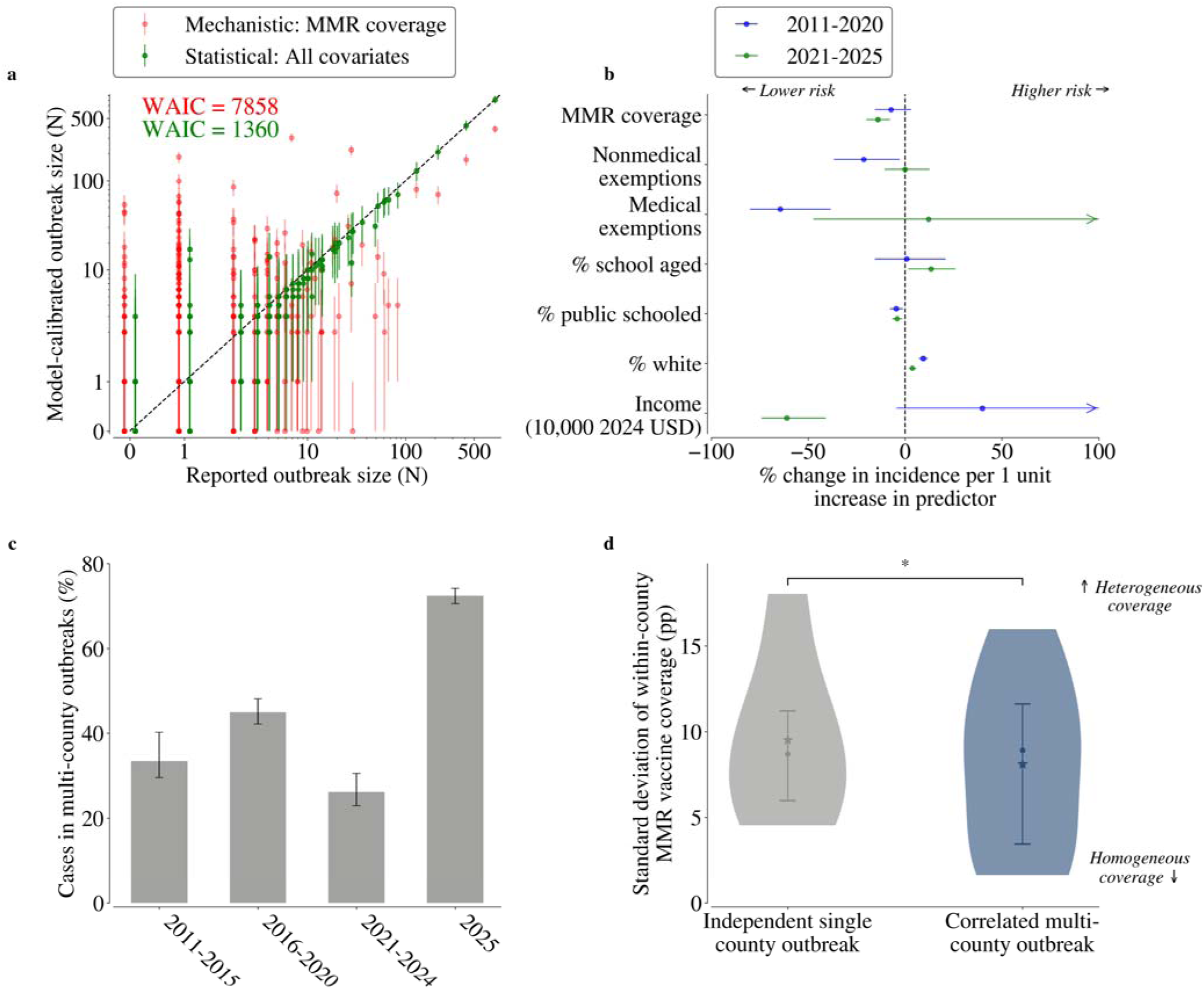
Calibrated model explains US measles cases at a county-level and reveals changing spatial patterns and correlates of outbreaks. (a) Calibration model performance estimating measles cases for all county-years reporting cases from 2021-2025. Green points show the posterior median estimates from a hierarchical Bayesian Poisson regression model based on MMR vaccine coverage and sociodemographic covariates, state and year fixed effects, and a conditional autoregressive spatial random effect at the county level. Red points show the median estimate from a final outbreak size mechanistic model based on county-level vaccine coverage. The WAIC refers to the Widely-Applicable Information Criterion, reported in the corresponding color for each model, where a lower value is better and the score penalizes model complexity. For both models, error bars show the 95% credible intervals. Model performance on out-of-sample validation data is shown in Table 1 and Figure 4. (b) Estimated relationships between outbreak size, vaccine coverage and exemptions, and socioeconomic and demographic covariates that had ≥95% posterior probability of being non-zero for the 2021-2025 time period. Points indicate the posterior median estimate, and error bars show the 95% credible interval. Credible intervals that extend beyond a 100% change in incidence were truncated for visualization: see Figure S8d for full credible intervals. (c) Spatial clustering of measles cases over time. We calculated the proportion of measles cases in counties that had at least three non-imported cases where a neighboring county also had at least three non-imported cases in the same year (“multi-county outbreaks”; y-axis) across time periods (x-axis). Error bars show 95% credible intervals from a beta sampling distribution. (d) Multi-county outbreaks have more homogeneous vaccine coverage. We calculated the unweighted standard-deviation of school district vaccine coverage for county-years when such data was available (data only available in 2024-2025; y-axis). We stratified counties by whether there was a ≥95% posterior probability that neighboring counties were correlated as a multi-county outbreak (blue), or not, which included counties with no outbreaks in neighboring counties (grey). Points show the median standard deviation across observations in each category, stars show the mean, and error bars show the 25^th^ and 75^th^ percentiles. The asterisk indicates that as the standard deviation of within-county MMR vaccine coverage decreased (more homogeneous susceptibility), the association between cases across neighboring counties increased significantly (*p* = 0.04), suggesting that evenly distributed measles susceptibility across a population increased the chance of an outbreak spilling across borders compared to isolated single county outbreaks which had larger within-county variation in vaccination.

**Figure 4:**
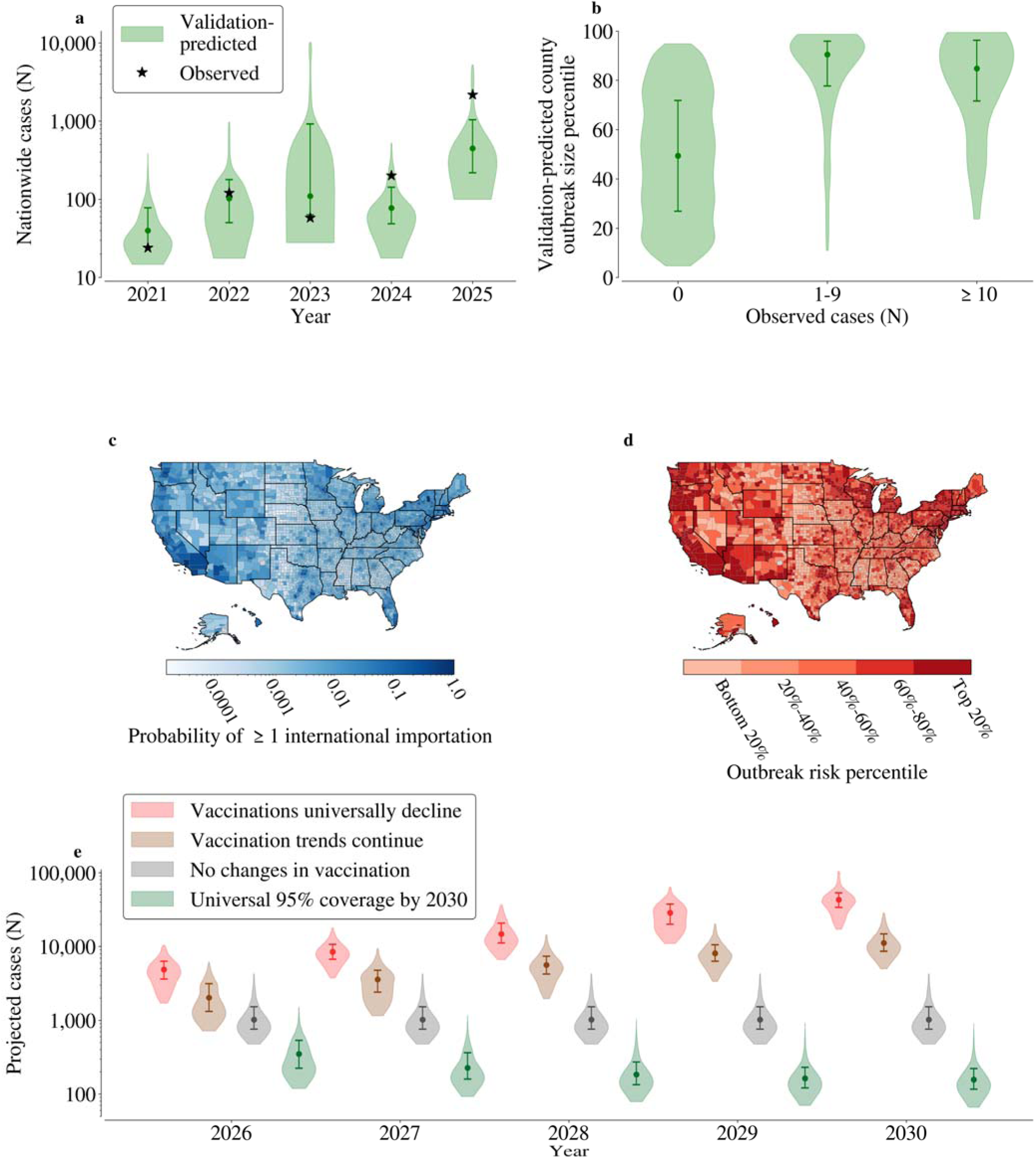
Spatial heterogeneity in county-level measles outbreak risk across the United States and measles projections over time using a validated model. (a) Validation performance of a statistical model that forecasted measles cases from 2021-2025 using a rolling window out-of-sample validation set. Violin plots show the 95% posterior range of forecasted annual nationwide measles cases (including importations) from the model when calibrated to all data prior to the validation year, points show the median, and error bars show the middle 50% posterior range. Stars show the observed number of reported measles cases. Model selection prioritized ranking county-level measles outbreak risk rather than projecting nationwide cases. (b) Validation performance of county-level measles risk percentiles from the statistical model (y-axis) compared to the out-of-sample validation data of observed county-level total measles cases (x-axis) from 2021-2025. The model predicted a higher risk percentile for counties with subsequently observed measles cases. (c) County-level map of the estimated probability of there being at least one international travel-associated measles importation over a year (See Figure S13 for independent validation of the importation process model). (d) County-level map of projected measles outbreak risk (based on predicted incidence) in the United States based on 2025 data, accounting for the risk of measles importations. Map shows substantial geographic heterogeneity in risk. (e) Projected incident annual nationwide measles cases 2026-2030 under various vaccination scenarios, using a log axis. “No change” refers to vaccine coverage not changing from 2025 data (or last available data). “Vaccine trends continue” refers to continuation of the trends presented in Figure 1d. “Vaccination trends universally decline” refers to switching counties whose coverage is increasing to instead decline at the national average and doubling the declines in all counties experiencing declines. “Universal 95% coverage” by 2030 refers to all counties with coverage below 95% experiencing a linear increase in coverage to reach 95% by 2030.

**Table 1:** Out-of-sample validation performance of models in predicting county-level measles cases in the United States for 2021-2025 when trained on data from preceding years.

| Model | Tested on | Sensitivity, 10+ cases (%) | Specificity, 10+ cases (%) | Positive predictive value, 10+ cases (%)<sup>3</sup> | Negative predictive value, 10+ cases (%) | RMSE (cases) |
| --- | --- | --- | --- | --- | --- | --- |
| Full spatial statistical model<sup>1</sup> | All cases in all counties | 94.9<br>[74.5, 100.0] | 82.9<br>[64.2, 97.6] | 4.1<br>[2.7, 8.2] | 99.2<br>[99.0, 99.4] | 34.6<br>[6.7, 103.9] |
|  | Outbreak cases only<sup>2</sup> | 78.3<br>[66.7, 100.0] | 74.9<br>[54.2, 91.8] | 2.9<br>[2.2, 3.7] | 99.3<br>[99.1, 99.4] | 42.8<br>[6.5, 140.1] |
| Mechanistic model | All cases in all counties | 93.1<br>[65.5, 100.0] | 82.8<br>[65.6, 97.3] | 4.1<br>[2.6, 7.3] | 99.2<br>[99.1, 99.5] | 33.8<br>[5.9, 104.1] |
|  | Outbreak cases only | 53.0<br>[33.3, 100.0] | 61.4<br>[42.1, 75.9] | 1.3<br>[1.1, 1.6] | 98.8<br>[98.3, 99.2] | 36.2<br>[6.1, 113.7] |

Selected model performance metrics are shown for top performing models (all models and additional model performance metrics shown in Supplementary Data S1-S3), determined through a model selection procedure that selects the models that best predict measles cases nationwide and classify counties with $\geq 10$ cases. Model metrics are calculated by averaging performance on annual predictions from 2021-2025, and the uncertainty intervals show the range across 2021-2025. Because many outbreaks are small, the root mean square error (RMSE) is calculated when there are at least 3 cases to assess performance in predicting larger outbreaks, which results in larger RMSE values. Other case cutoffs for the RMSE are available in Supplementary Data S3, including other metrics such as the Weighted Interval Score, Continuous Ranked Probability Score, and relative Mean Absolute Error for model comparisons.
<sup>1</sup> Includes all covariates and partially-pooled state-specific coefficients for vaccine coverage and exemption prevalence.
<sup>2</sup> Tested on total cases minus confirmed importations in counties with at least one measles case (i.e., an importation).
<sup>3</sup> The prevalence of counties with 10+ cases was 0.5% (0.06, 1.7).

We found that recent outbreaks were considerably more spatially clustered. We identified multi-county outbreaks by searching for counties that had at least three outbreak cases (the minimum number of cases required to be considered a CDC-level outbreak) whose neighbors also had at least three outbreak cases in the same year. The percentage of measles cases in multi-county outbreaks increased from 34.8% (32.7, 37.8) in 2011-2024 to 72.3% (70.5, 74.2) in 2025 (Figure 3c). From 2021-2025, the Bayesian Poisson model estimated that cases between neighboring counties in multi-county outbreaks had a residual correlation of 0.89 (0.80, 0.94) compared to 0.37 (–0.48, 0.80) from 2011-2020 (Figure S11), indicating that outbreaks from 2021-2025 that were larger than expected from vaccine data and covariates alone were spatially clustered and suggestive of transmission across counties that has not occurred previously. Moreover, as outbreak size increases across neighboring counties, so too does the relationship between vaccine coverage and outbreak size (Figure S11), and sensitivity analyses indicated that this remained even when accounting for similarities in vaccine coverage across neighboring counties (Figure S6c).

We quantified predictors of spatially clustered outbreaks to explain their prominence in recent measles outbreaks. We hypothesized that large outbreaks confined to a single county reflected high variability in within-county MMR vaccine coverage, indicating transmission among tightly knit under-immunized communities, whereas spatially clustered multi-county outbreaks arose from consistently low (i.e., more homogeneous) within-county MMR vaccine coverage that led to more widespread transmission across counties. We obtained MMR vaccine coverage for school districts within a county (available for 126 county-years in 2024-2025, representing 40.7% of measles cases during this time period) and calculated the standard deviation of within-county MMR vaccine coverage (Figure S12). Lower standard deviations indicated more homogeneous MMR vaccine coverage, showing more uniform susceptibility across the county rather than localized within specific communities. Focusing on larger outbreaks (at least 10 cases) where transmission across counties is more relevant for public health response, we found that homogeneity in within-county MMR vaccine coverage was significantly related to spatially clustered multi-county outbreaks (Figure 3d; Table S1) while heterogeneity in within-county MMR vaccine coverage predicted independent single county outbreaks, supporting the hypothesis that recent multi-county measles outbreaks are reflective of uniformly-low vaccine coverage that enables transmission throughout the county and across borders. Sensitivity analyses indicated that this relationship existed across all outbreak sizes and strengthened as outbreak size increased, was consistent across states, and was robust to the proportion of public schools in the county (Table S1).

### Validated spatial statistical model predicts county-level measles cases

We evaluated the validation performance of the hierarchical Bayesian Poisson regression models in predicting county-level measles risk compared to the benchmark mechanistic model using rolling validation where models were calibrated to outbreaks occurring before each prediction year from 2021-2025. Because these models predicted locally acquired cases conditioned on an internationally imported index case in the county, we developed a stochastic model of the measles importation process, so we could predict total expected county-level measles cases nationwide. Calibrated to data from 2011-2020, the importation model predicted counties from 2021-2025 with at least one imported measles case with an area under the receiver operating characteristics curve of 0.92 (Figure S13 shows a precision-recall curve).

We joined the predictions from the importation model with estimates of locally acquired cases from the outbreak models to predict county-level risk nationwide. We found that models that included spatial relationships between counties consistently performed better in predicting county-level measles cases than those that did not (Supplementary Data S1-S3). In particular, the Bayesian Poisson statistical model with all covariates and a conditional autoregressive spatial term demonstrated high concordance with predicted measles cases from 2021-2025 (Figure 4a) and high sensitivity and specificity in classifying counties with at least 10 cases (Table 1; Figure 4b). While both the statistical and mechanistic (which reflects rank-ordering counties by vaccine coverage alone) models had comparable performance predicting which counties would have at least 10 cases (Table 1), the full spatial statistical model with all covariates and a conditional autoregressive spatial term was better at classifying counties by outbreak (local transmission) cases, which are most relevant for re-emergence risk and public health intervention, and this was consistent across outbreak sizes (Supplementary Data S2).

Consequently, we selected this model as our outbreak model for subsequent analyses, though many models performed well. With this model, predicted outbreak size percentiles by year were directly related to observed outbreak sizes from 2021-2025 (Figure 4b), and the model reproduced the outbreak’s size percentile and the overall outbreak size distribution (Figure S14), supporting its ability to prospectively differentiate higher and lower risk counties prior to outbreaks to enable pro-active public health intervention.

### Near-term measles projections in the United States find that existing vaccine declines may lead to widespread transmission

Using our validated prediction model, we projected measles re-emergence risk nationwide and under future vaccine coverage scenarios. We found that measles risk is spatially concentrated and may worsen significantly if current vaccination declines continue. We predicted the probability of an international travel-associated measles case by county (Figure 4c) and combined those predictions with our full spatial statistical model to project measles risk at the county-level nationwide (Figure 4d). We find that outbreak risk is both spatially concentrated and geographically widespread across the entire US. The model-predicted risk map highlights areas that have documented recent outbreaks while identifying new regions that may be at risk in the future. To better understand future risk, we projected measles cases from 2026-2030 under four scenarios: if vaccine coverage stays the same as 2025, vaccination trends continue (as described in Figure 1d), vaccination declines accelerate twofold, and all counties achieve 95% MMR vaccine coverage by 2030 (Figure 4e). If vaccination trends continue, our analysis predicted 30,700 (12,500, 73,300) cases (Table S2 presents the impact of model priors on these estimates) by 2030. Comparatively, if declines accelerate twofold, that is projected to increase to 100,700 (41,500, 215,400) cases, and by contrast, if all counties reach 95% MMR vaccine coverage by 2030, that would be reduced to 1,100 (500, 5,200) cases. Nevertheless, measles cases remain concentrated in a subset of counties: across all vaccine scenarios (Figure S15), we estimated that greater than 90% of cases would be in the 10% of counties with the highest model-predicted risk (Supplementary Data S4; Figure S14).

### Spatially targeted interventions may mitigate measles re-emergence nationwide

We evaluated the impact of increasing vaccine coverage to mitigate measles re-emergence risk in the US, such as removing nonmedical exemptions to childhood vaccine requirements. Nonmedical exemption prevalence varies widely and averages 4.3% in US states where they are allowed (Figure 5a). We analyzed changes in observed county-level MMR vaccine coverage from historic state-level policies that have removed nonmedical exemptions (California, Connecticut, Maine, and New York) to quantify the historic impact of such policies and reasonable increases in vaccine coverage from interventions (Figure 5b). We found that county-level MMR vaccine coverage following policy enactment increased by 0.73 (0.61, 0.82) percentage points per nonmedical exemption percentage point prior to policy enactment, corresponding to an average of a 2.3 percentage point increase in county-level MMR vaccine coverage across all counties in states where such polices were implemented with larger increases in counties with lower vaccine coverage (Figure 5b). This estimate was consistent across all states implementing such policies, and coverage remained elevated for at least two years after the policy was enacted (Table S3).

**Figure 5:**
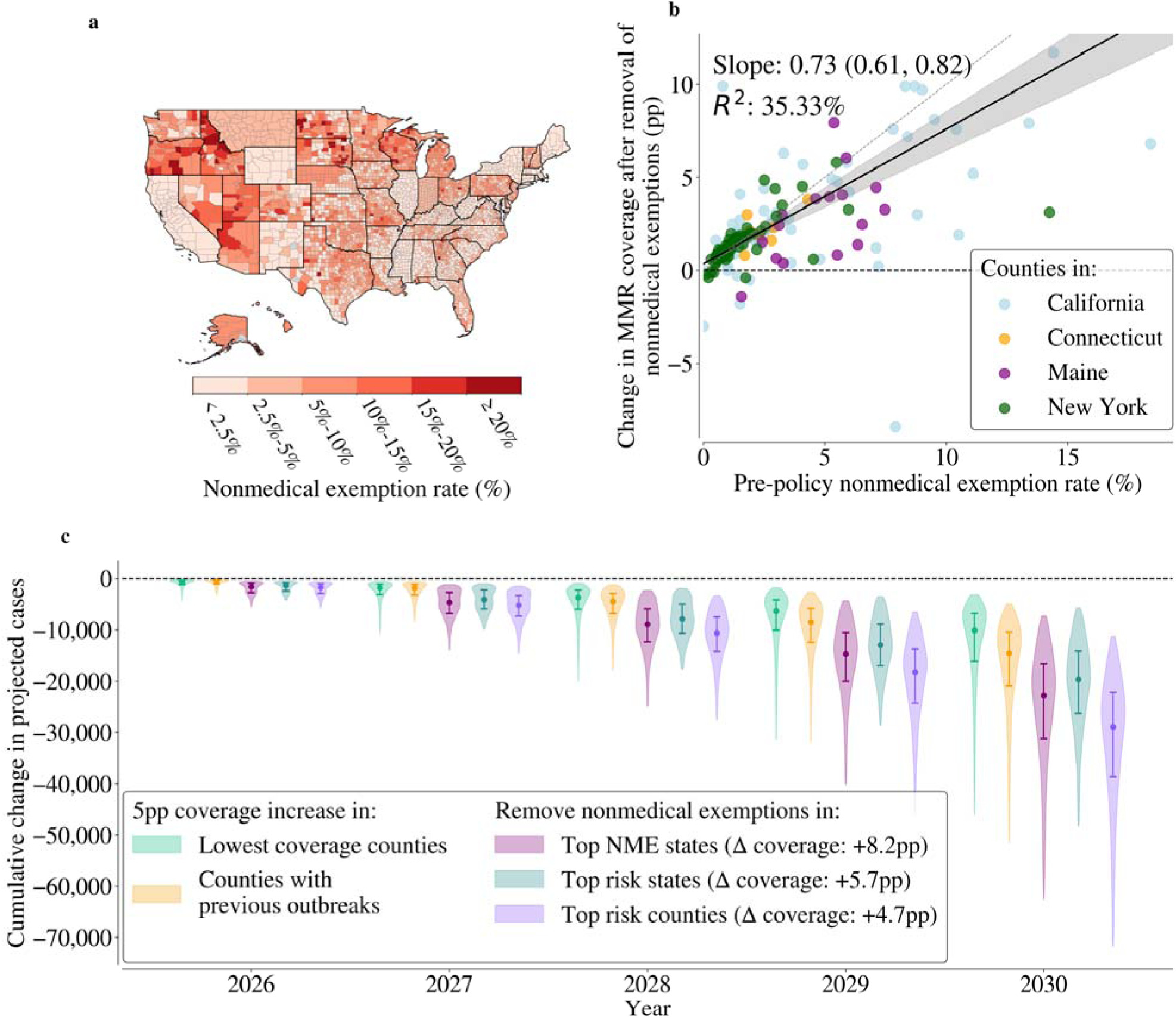
Public health impact of targeted vaccine exemption policy changes and increased vaccination to mitigate measles re-emergence in the United States. (a) County-level prevalence of nonmedical exemptions to school vaccine requirements (mostly among entering kindergartners) from the most recent year of available data, mainly 2024-2025. Source data are from Fattah, Stoffel et al., 2026.^44^ (b) Observed change in county-level MMR coverage after state policies removing nonmedical vaccine exemptions were enacted. Points show the observed percentage point (pp) change in a county’s vaccine coverage one year after the policy (y-axis) as a function of its nonmedical exemption prevalence prior to the policy being enacted (x-axis). The dashed grey line shows the theoretical line that indicates that all individuals with nonmedical exemptions subsequently became vaccinated. The grey line shows a Theil-Sen robust regression line, and the grey shaded area shows the 95% confidence interval around the regression line. In some cases, policies were enacted alongside public health efforts to increase vaccination. (c) Projected impact of various strategies to increase vaccine coverage over time. We tested five strategies: two where vaccine coverage was increased directly and three where we removed nonmedical exemptions. In particular, we modeled increasing vaccine coverage by 5 percentage points (pp) in (i) counties in the bottom decile of vaccine coverage and (ii) counties with previous outbreaks from 2011-2025. We also evaluated removing nonmedical exemptions in (iii) the five states with the highest nonmedical exemption prevalence, (iv) the five states with the highest model-predicted outbreak risk, and (v) the top 10% of counties with the highest model-predicted outbreak risk. Violins show the 95% posterior range of cumulative change in cases over time, points indicate the median, and error bars show the middle 50% posterior range. Projections are made assuming that vaccination trends continue. See Figure S16 and Supplementary Data S5-S6 for sensitivity analyses under different future vaccination scenarios.

We evaluated where the expansion of such policies and vaccination should be prioritized. We modeled five strategies: three where nonmedical exemptions were removed to increase vaccine coverage and two where vaccine coverage was increased by 5 percentage points directly through public health outreach efforts (an increase in vaccine coverage similar to that from removal of nonmedical exemptions in the targeted locations). We found that spatially targeted interventions in the counties of highest model-predicted risk were most impactful while intervening in counties based on vaccine coverage alone or previous outbreaks in the county was least impactful. In particular, targeted efforts to increase vaccine coverage in the top decile of counties with the highest model-predicted measles outbreak risk reduced projected national cases to 2030 by 93.3% (81.3, 97.9) and required increasing vaccine coverage the least by an average of 4.7 percentage points (Figure 5c). In alternative setups of this analysis that include model prediction errors, we consistently find that intervening in the top 20% of counties by model-predicted risk reduces at least 80% of cases (Figure S14). Intervening in the five states with the highest model-predicted outbreak risk versus in the five states with the highest nonmedical exemption prevalence had similar projected reductions in measles cases to one another (64.9% (26.8, 89.0) vs 75.0% (41.2, 92.8)), though targeting high exemption states required a larger increase in vaccine coverage (8.2 percentage points) compared to targeting high risk states (5.7 percentage points). Across varying scenarios of changes in vaccine coverage to 2030, we robustly found that intervening in the counties of highest model-predicted risk reduced re-emergence risk the most (Figure S16; Supplementary Data S5-S6). Consequently, our results indicate that spatially targeted interventions reduce measles cases the most, and certain state-level interventions provide moderately smaller reductions.

## Discussion

In this study, we found that measles re-emergence risk is spatially concentrated while geographically distributed across the United States, and prioritizing interventions in the highest risk locations may effectively mitigate measles re-emergence nationwide. Using an assembled dataset of county-level annual measles cases and MMR vaccine coverage and exemptions data in the US from 2011-2025, we found evidence for a significant epidemiologic shift indicative of measles re-emergence in the US and that outbreaks have become increasingly explainable and spatially clustered over time. We synthesized these insights into a validated county-level predictive model of measles that accurately stratified counties based on outbreak size from 2021-2025 and projects cases under future vaccination scenarios, demonstrating that measles cases may grow significantly if current vaccination declines continue. Most notably, we found that prioritizing modest (5 percentage point) increases in vaccine coverage in the 10% of US counties predicted to be highest risk for large outbreaks may effectively reduce re-emergence nationwide, and using model-based risk identification was more impactful than conventional approaches such as targeting based on low vaccine coverage alone. These results have broad implications for control of measles in the US and other settings with re-emerging measles amid declining vaccination.

Our study presents evidence for an epidemiologic shift associated with measles re-emergence in the US. The spatially fine and temporally long scale of the measles data used in this analysis enabled characterizing measles epidemiology over the elimination era, through the COVID-19 pandemic, and to the recent period of sustained outbreaks threatening elimination status, enhancing comparisons across different time periods. We found that the relationship between county-level vaccine coverage and outbreak size has strengthened significantly in recent years, as multi-county outbreaks fueled by counties with homogeneously low vaccine coverage have become increasingly prominent. Historically, measles outbreaks in settings with elimination have been among small, tightly knit, socially clustered communities that are underimmunized,^9^ and county-level measures of vaccine coverage often do not properly reflect such clusters.^34^ Instead, a strengthening relationship between county-level vaccine coverage and outbreak size along with increasing spatial clustering of multi-county outbreaks suggests that measles transmission has become more broadly sustained across the general population, driven by widespread declines in vaccine coverage contributing to more homogeneously low vaccine coverage. This interpretation is further supported by analyses of sub-county vaccination coverage where counties with heterogenous vaccine coverage (i.e., clustered communities) were more likely to stay as single county outbreaks whereas multi-county outbreaks demonstrate more homogenous vaccine coverage that enables the spread of measles throughout the general population. Together, these data underscore that shifts in measles epidemiology in the US indicate re-emergence patterns with increasingly widespread transmission throughout counties and across county borders.

We found that measles cases are highly spatially concentrated in a small subset of counties across the US. While cases are rising nationwide, the largest outbreaks that disproportionately drive total cases occur in a limited set of counties. Identification of these counties is improved with accounting for factors beyond vaccine coverage alone, such as sociodemographic covariates and the spatial relationships between counties. Models that included these factors showed better predictive performance in classifying counties by local outbreak size. This indicates that once vaccine coverage is low, additional covariates that reflect population structure, mobility, and public health response are necessary to accurately characterize measles re-emergence risk.

Because re-emergence risk is spatially concentrated in a subset of counties, targeted preventive efforts to increase MMR vaccine coverage in those counties may effectively mitigate measles cases nationwide. We demonstrated that policies that have removed nonmedical exemptions from childhood vaccines have been associated with significant increases in county-level MMR vaccine coverage, consistent with prior literature,^37^ and these gains are consistent across different states and sustained over time; these policies may be implemented alongside other efforts to increase vaccine coverage,^36^ such as school outreach,^46^ catch-up community-level vaccination campaigns,^47^ and provider-level reminders.^48^ While nonmedical exemption policies are often state-level, this analysis found that targeting public health measures with similar impacts on increasing vaccination at a finer spatial scale to the counties at greatest model-predicted risk was more impactful than policies implemented at the state level, underscoring the need for spatially resolved estimates of measles risk and fine-scale targeting measures.

Our spatial statistical model had high performance in predicting measles outbreaks, suggesting a future role in proactive public health response to identify counties at risk and prioritize intervention efforts. We conducted rigorous forward-looking validation analyses to predict county-level measles cases across all counties in the US from 2021-2025; such validation exercises are relatively uncommon beyond short-term forecasting,^49^ and doing so improves confidence in the validity of our projections.^50^ Most importantly, the model performed well at categorizing counties without outbreaks as low-risk and consistently placing counties with outbreaks at high risk percentiles, which is more important for pre-emptive public health response than projecting total national annual cases. The PPV was modest because large outbreaks remain nationally infrequent. Moreover, using this model to identify areas for targeted interventions substantially reduced projected measles re-emergence risk. Nevertheless, it remained challenging to forecast the absolute number of cases in large outbreaks, underscoring the transmissibility and stochasticity of measles and that models should be complemented by active surveillance when being used in real-time public health response.

Our international importation model presents an alternative approach to predicting county-level international travel-acquired measles cases. Previous methods often use origin-destination air travel data, measles incidence in countries around the world, and MMR vaccine coverage in the domestic destination county. These approaches have demonstrated moderate performance in predicting measles importations,^42,43^ but they require significant costly ancillary data. Our approach relied solely on historical publicly reported state-level measles importations, which we modeled as a stochastic process with county assignment based on population, achieving strong validation performance compared to existing approaches.^42^ However, our model assumed that annual importations remain consistent over time, and as the global incidence of measles rises, it is likely that international measles importations may rise and shift geographically.

Our study has limitations. First, our underlying data had limitations. Although we assembled a county-level annual measles case count dataset for this study, we did not have access to individual-level case data or relevant information to determine who infected whom; therefore, we could not confirm that cases in a county-year were epidemiologically linked or if the same outbreak spanned multiple years or counties. Despite these challenges, our models had high validation performance using conventionally reported case data, which would be the primary data source available in practice. Second, our county-level MMR vaccine coverage data had differences across states in the definition of MMR vaccination (versus up-to-date vaccination) and the age groups included (entering kindergarteners versus entering K-5 schoolchildren), which we addressed by using state-fixed effects and allowing for state-level variance in the relationship between vaccine data and outbreak size. Some counties and years had missing data, especially among older time periods. Moreover, these data also represent only a sample of a county’s actual measles immunity as older children, adolescents, and adults also contribute meaningfully to population immunity but were not fully captured in this study. However, our results suggest that vaccine coverage in these age groups is related to both younger and older age groups and therefore may be a more representative measure. There is also likely limited variation in immunity in older adult populations.^51^ Additionally, while we analyzed school district level variation in vaccination in a sub-analysis, such a dataset for both cases and vaccination were not available over the entire study period. Higher resolution school-level data can help pre-emptively indicate hotspots of re-emergence risk otherwise obscured by county-level data,^34^ and our study supports the finding of spatial variability in re-emergence risk by alternatively demonstrating empirical evidence of re-emergence using historical county-level measles cases and further indicating through intervention scenarios that spatial targeting can help optimize public health response. Finally, our importation dataset was incomplete. We could only identify county-level cases that were internationally imported in a subset of the dataset where they could be unambiguously assigned to a county from the reported state-level data. Nevertheless, our importation model showed high out-of-sample validation performance across all counties with these data. Similarly, we did not explicitly model domestic importations between counties.

Our modeling methods had additional limitations. First, our hierarchical Bayesian Poisson regressions could not conclusively explain the mechanism behind the epidemiologic shifts, though we conducted sensitivity analyses to ensure our findings were robust to variations in the underlying data and model parameterizations. Second, although our results indicated that measles outbreaks are becoming increasingly spatially clustered over time, it remained challenging to disentangle whether this could have instead been from unobserved similarities between counties (such as shared public health resources, schools, or persistent infection exposure) or epidemiologically separate outbreaks. Finally, while our approach identifies epidemiologic shifts, our predictions are trained on recent measles epidemiology, which may continue to shift as re-emergence occurs. This model is designed for elimination and peri-endemicity settings but is not optimally designed to model endemic transmission. A discussion of additional limitations is available in the Appendix.

We demonstrate that measles epidemiology has shifted as the US observes sustained and widespread transmission. Outbreaks have become increasingly explainable, and predictive models can be a valuable tool to identify the areas at greatest risk, particularly as vaccination rates decline and outbreaks are anticipated to grow. Moreover, we found that outbreak risk is spatially concentrated, and spatially targeted interventions that increase MMR vaccine coverage in the subset of locations at highest risk may significantly reduce national risk, a pattern that persists even if vaccination rates decline further. As childhood vaccination rates decline globally, our findings underscore that predictive models and spatially targeted interventions can significantly reduce outbreak risk to mitigate the re-emergence of vaccine-eliminated diseases.

## Methods

### County-level measles case and vaccination data

We assembled annual county-level MMR vaccine coverage and vaccine exemption^44,45^ and measles case count data from 2011-2025 for the United States (see Appendix). These data were predominately obtained from state health and education departments through direct outreach or publicly available sources. MMR vaccination coverage was defined as the percentage of entering school children (mainly kindergarteners) in each county who had received two-doses of MMR vaccine or alternatively had received all required state-level vaccines and doses if coverage for MMR alone was not available. Additional details on state– or year-specific differences in these data is provided in the Appendix. Measles cases were defined as a lab-confirmed case of measles reported to the state public health department, with an additional step to map them to a known county or county-equivalent units (see Appendix). To differentiate between local and imported cases, state-level annual importation data were obtained from the US Centers for Disease Control and Prevention with assignment to the underlying county when possible (see Appendix). Additional data processing decisions were made, resulting in the removal of three county-years of outbreaks (see Appendix). We also obtained county-level sociodemographic data mainly from the five-year American Community Survey;^52^ variables included the fraction of the population under the age of five, the fraction of the population that is school-aged (under the age of 18), the fraction of the population enrolled in school who are in public school, the average and standard deviation of household sizes across the county, the fraction of the population that is white, the fraction of the population with a graduate degree, the median income, and the population density. This study was approved by the Stanford University Institutional Review Board.

### Model overview

We developed a two-part modeling framework to predict annual county-level measles cases in the US. First, the “importation model” estimated the number of annual county-level imported measles case due to international travel, *I,* based on a stochastic model that describes measles importations. Second, the “outbreak model” estimated the total number of locally acquired measles cases, *C*, given an internationally imported case. We developed multiple versions of the outbreak model, including mechanistic and statistical models that each predicted local transmission case counts. The mechanistic model depended only on MMR vaccine coverage while the statistical model included covariates such as vaccine coverage, vaccine exemptions, county-level sociodemographic variables, and spatial effects. All models were independently calibrated and validated. To obtain the expected number of annual measles cases in a US county, *T,* we used the following formula:

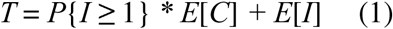

### Importation model

We modeled the number of county-level internationally imported measles cases (arising from an international traveler who was exposed to the virus in another country and then returned to the US) using Bayesian Poisson-Gamma (negative binomial) processes. Since the US CDC reports importation data aggregated by state, we began by independently describing empirical negative binomial processes for the number of observed yearly statewide imports from 2011 to 2025. To ensure meaningful estimates even for states that never had an international importation during the study period, we assigned a Uniform(0, 1) prior distribution on the probability of a state having at least one import in a year. Thus, the average number of annual importations in a state μ*s* is given as follows:

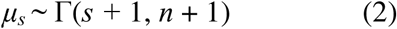

where Γ is a gamma distribution in shape-rate form, *s* is the total number of imports over the *n* years of data (in this case, from 2011-2025), and the addition of unity to each of the gamma distribution parameters reflects the uniform prior. To estimate each county’s imports, we distributed state-level imports to counties by population size. We validated this approach by comparing to the subset of county-years with known import data, described below. We calculated both the probability that the county would have at least one international import, *P*{*I* ≥ 1}, and the total number of international imports, *E*[*I*].

### Outbreak model

We modeled the number of annual locally acquired measles cases (outbreak size) given an international import using a benchmark mechanistic model that depended on measles *R_0_* (assigned as 12 based on the literature^12^) and MMR vaccine coverage alone and a hierarchical Bayesian Poisson regression (statistical) model that optionally included additional covariates. Our benchmark (base case comparison) mechanistic model estimated annual county-level local measles cases using the final outbreak size of a homogeneous susceptible-infected-recovered (SIR) compartmental model with an initial susceptible population of people who did not have all-or-nothing vaccine-derived immunity (See Appendix for further details). In analyses that rank order counties, this benchmark model ranks counties by vaccine coverage, such that model formulation assumptions (such as *R_0_*) do not impact ranking. This formulation provides a nontrivial solution for the population infected during the outbreak when the susceptible population is greater than the herd immunity threshold,^53^ but outbreaks can still occur when the susceptible population is less. In this regime, we estimated outbreak size using a stuttering chains process.^54^ To account for state– and year-level differences in reporting and public health response, final outbreak size estimates in our inferential analyses were passed through a Poisson regression that incorporated state– and year-fixed effects. The mechanistic model assumed that all cases in a county-year were epidemiologically linked; as such, we conducted a sensitivity analysis assuming that each importation could lead to its own transmission chain (Supplementary Data S1).

The hierarchical Bayesian Poisson model regressed on county-level MMR vaccine coverage, nonmedical and medical vaccine exemption prevalence, and sociodemographic variables. To account for state-specific differences in vaccine coverage and reporting over time, the model included state– and year-fixed effects. We also allowed the relationship between outbreak size and MMR coverage, nonmedical exemptions, and medical exemptions to vary by state via partial pooling to further consider state-level differences in vaccine reporting and the nonlinearity between vaccine measures and outbreak size. Finally, to model relationships such as outbreak spillovers between neighboring counties, we included a conditional autoregressive (CAR) random effect that captures unobserved spatial heterogeneities and dependencies between spatially adjacent (neighboring) counties. Our full model structure for predicting the number of measles cases given an import in county *c* in state *s* in year *y* was as follows:

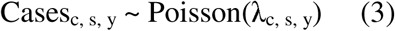

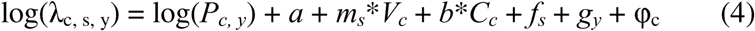

where *P_c,y_*is the population in the county for the given year, *a* is the log baseline risk of cases across all locations and years, *m_s_* is the state-specific set of hierarchically structured coefficients for MMR vaccine coverage, nonmedical exemptions, and medical exemptions that partially pool towards a set of nationwide coefficients via a normal distribution, *b* is the set of coefficients for county-level sociodemographic variables described above*, f_s_* and *g_y_* are the state– and year-fixed effects, respectively, and φ_c_ is the county’s spatially correlated random effect from the proper conditional autoregressive (CAR) process which estimated the residual spatial autocorrelation between neighboring county’s cases. In our analyses that evaluated the changing relationship between covariates and outbreak size continuously by year, we expanded this model to allow the coefficients for vaccine data and county-level variables to vary over time via a Gaussian random walk.

### Model calibration

We independently calibrated three models to the relevant measles data: the importation model was calibrated to annual state-level international measles imports, and the benchmark mechanistic final outbreak size model and hierarchical Bayesian Poisson regression model were calibrated to annual county-level locally acquired measles cases. For the inferential analyses, all models were calibrated to data from 2011-2025, which was broken into two time periods of 2011-2020 and 2021-2025 when explicitly evaluating differences over time.

Each model was calibrated using the relevant procedure, as described below. Since the posterior distribution for estimated state-level measles importations was analytically tractable, it was solved explicitly using Equation 2. Based on observed differences, both outbreak size models were calibrated separately to two time periods, 2011-2020 and 2021-2025. The Poisson regression of the output of the final outbreak size model was fit using maximum likelihood estimation. The hierarchical Bayesian Poisson regression was calibrated using No U Turn Sampler (NUTS) Markov chain Monte Carlo (MCMC) algorithm.^55^ Weakly informative or regularizing priors were chosen for all parameters; priors are described in full in Table S4.

### Model validation

We independently validated the importation model, the outbreak models, and the joint models by combining the importation model with each outbreak model according to Equation 1 in a forward-looking rolling window evaluation where we compared predicted and observed county-level cases from 2021-2025. In this validation, the model was trained only on data up to a given year (e.g., 2011-2020) and then evaluated on its performance in predicting cases in all counties in the following year (e.g., 2021). The importation model was assessed using the area under the receiver operating characteristics curve (AUC) and average precision (positive predictive value) in predicting whether a county would have at least one international importation, available for a subset of counties. We assessed the AUC, sensitivity, specificity, positive predictive value, and negative predictive value in forecasting whether a US county would have an outbreak of at least five, ten, or twenty cases along with the *R*^2^, Spearman correlation coefficient, root mean squared error, median absolute error, median fold error, the weighted interval score, the continuous ranked probability score, and the relative mean absolute error between predicted and observed cases across all counties with at least three, five, or ten cases and all counties with at least that many outbreak cases, and the absolute number of annual cases predicted across the entire US. We also assessed concordance between observed outbreak size and average predicted outbreak size percentile across all US counties.

### Near-term scenario analyses of future measles cases under various vaccination scenarios

We used the top performing model to project future measles cases to 2030 under four scenarios of changes in vaccinations informed by historic trends in county-level vaccinations, assuming as many measles importations as there were in 2025. In our main analysis, we evaluated four plausible scenarios: (i) no change in MMR vaccine coverage from 2025 data (or last available data), (ii) continued trends in MMR vaccine coverage based on 2021-2025 data, (iii) worsening decline in MMR vaccine coverage, with a doubling in the rate of decline in counties currently experiencing declines and counties with increasing MMR vaccine coverage switching to a decline at the historic national average, and (iv) universal 95% MMR vaccine coverage, with counties with MMR vaccine coverage below 95% experiencing a linear increase in MMR vaccine coverage to 95% by 2030 and counties with MMR vaccine coverage above 95% experiencing no changes.

### Intervention scenario analyses

We evaluated how targeted interventions increasing vaccine coverage may change future case projections. We grounded our analyses with empirical estimates on the impact of policies that remove nonmedical exemptions to childhood vaccine requirements. Using our county-level dataset of vaccine coverage, we calculated the change in county-level vaccine coverage associated with states that have passed policies removing nonmedical exemptions (California, Connecticut, Maine, and New York). We related nonmedical exemption prevalence prior to the policy with the change in vaccine coverage one year after policy was enacted and used a Theil-Sen robust regressor to estimate the relationship between the two.^56,57^ Applying this relationship, we estimated how removal of nonmedical exemptions would impact vaccine coverage, and then modeled expected measles cases under three scenarios of expanding these policies to (i) the five states with the highest nonmedical exemption prevalences, (ii) the five states with the highest model-predicted risk based on projected measles cases, and (iii) the top decile of counties with the highest model-predicted risk based on projected measles cases. Because the intervention scenario analyses tested the impacts of policy expansion, states or counties that had already removed nonmedical exemptions or had a nonmedical exemption prevalence of zero were omitted from consideration. We also considered two comparator strategies where we increased vaccine coverage by five percentage points in (i) the bottom decile of counties by vaccine coverage, and (ii) counties that previously had at least three cases in a year anytime from 2011-2025. We calculated the population impacted by each of these policies and estimated the number of vaccine doses given under each strategy.

### Sensitivity analyses

We conducted numerous sensitivity analyses to evaluate the robustness of our description of the epidemiologic shift in measles outbreaks and evaluation of what kinds of interventions were most impactful in reducing re-emergence risk. Further details are available in the Appendix. In brief, first, we repeated our analyses using the population-weighted five-year average of vaccine coverage and vaccine exemptions. Second, we refit each of the models using various approaches for controlling for a county having previous outbreaks. Third, we repeated our analysis for how the relationship between vaccine coverage and outbreak size has changed over time under various parameterizations of allowing the relationship between control covariates, spatial relationships, and outbreak size to change over time. Fourth, we estimated the impact of policy interventions under different scenarios of future vaccination coverage and assumptions about reasonable changes in vaccine coverage from policy response.

### Data sharing statement

Study data that are not considered identifiable are available online in the GitHub repository. Requests for measles case data, which are not publicly posted due to potential identifiability concerns, can be obtained from the authors.

### Code sharing statement

All code is available online under an MIT GNU public license.^58^

## Data Availability

https://github.com/chiragkumar9/measles-takeoffs

## Acknowledgments

We acknowledge the state health and education departments that provided data for this study. We acknowledge input from news team members from NBC News in collection and processing of the vaccination dataset. The study content is solely the responsibility of the authors and does not necessarily represent the official views of the California Department of Public Health, California Health and Human Services Agency, National Science Foundation, or National Institutes of Health.

## Author contributions

Mr. Chirag K. Kumar and Dr. Nathan C. Lo had full access to all the data in the study and take responsibility for the integrity of the data and the accuracy of the data analysis.

Study concept and design: CKK, NCL

Statistical analysis: CKK, NCL

Analytic coding: CKK

Acquisition, analysis, or interpretation of data: All authors

First draft of the manuscript: CKK, NCL

Critical revision of the manuscript: All authors

Contributed intellectual material and approved final draft: All authors

## AI statement

Claude LLMs (Sonnet 4.5-4.6) were used for additional code review of human-written analytic code. OpenAI LLMs (ChatGPT 5.2) were used solely for minor editorial assistance (e.g., grammar, clarity, and formatting). All ideas, analysis, writing, interpretation, coding, and conclusions are the original work of the authors, who retain full responsibility for the content.

## Supplementary Materials

Materials and Methods

Figures S1 to S14

Tables S1 to S4

Descriptions of Supplementary Data S1 to S6

References (#1-#27)

