## Supplemental Appendix for "Epidemiologic shifts predict spatial patterns of measles re-emergence in the United States"

**Supplementary Methods**
**Supplementary Figures**
**Supplementary Tables**
**Supplementary References**

### Supplementary Methods

#### Additional information on data acquisition and processing

*County-level measles cases*

Total annual county-level measles case count data were gathered for 2011-2025 following a consistent process across all states and years. We began with annual lab-confirmed state-level measles case count data as reported to the Nationally Notifiable Diseases Surveillance System of the United States Centers for Disease Control and Prevention (CDC); this source reported both annual internationally imported and locally acquired cases.[1] From this, we assigned total annual state-level cases to counties using a two-step process. For most state-years (76%), the relevant health department in the state provided county-level assignments of measles cases either through publicly available health dashboards, reports, or record requests. If the total number of annual state-level measles cases reported by the CDC and the state differed, we defaulted to the state data (these differences totaled 0.76% of cases; the maximum difference in a single state-year was 17 cases from California in 2014 when a significant outbreak occurred in Disneyland that included residents from other states). For the subset of states where such data were not available (21.5% of total cases), we used news and public health press releases that assigned measles cases to a county. Such reports had to explicitly note that the case was confirmed by the county or state health department as being in that county. Probable cases and cases among non-residents of a county were not included. Over the entire study period, we mapped 96.5% of CDC-reported measles cases to their underlying county or county-equivalent within the state.

There were additional data processing decisions. In our inferential calibration analyses, we included cases until the end of January 2026 in the 2025 case counts for counties in South Carolina only. This was because South Carolina only reported cases at the spatial resolution of regions rather than counties for 2025 alone, but 2025-2026 cumulative cases (which included a notable outbreak centered in Spartanburg County) were available at the county-level. In our validation framework, when predicting 2025 cases for South Carolina, we did not include 2026 cases in the observed value when comparing to our predictions. We also removed county-years that contained outbreaks that occurred among refugee populations or migrants that had recently arrived in the United States from analyses since these populations could not have been captured in the relevant count-level vaccine data. This consisted of three county-years: (i) Cook County, Illinois in 2024 (Venezuelan migrants);[2] (ii) Monroe County, Wisconsin in 2021 (Afghan refugees);[3] and (iii) Pinal County, Arizona in 2016 (US Immigration and Customs Enforcement detention center outbreak).[4]

*County-assignments for state-level reported internationally imported/travel acquired measles cases*

Measles cases in a county-year could either be internationally imported or locally acquired. An internationally imported measles case was defined as a measles infection that resulted from international travel and exposure to measles virus outside the US (based on an exposure window) where the individual then developed a measles case within a set time upon returning to the US and were diagnosed [5]. Comparatively, a locally acquired measles case was defined as any measles case not classified as an internationally imported case, indicating that the infected individual had no history of recent international travel. Thus, locally acquired measles cases may have included domestically imported cases (i.e., a patient acquiring measles in a different county or state than their residence and returning to their residence county for diagnosis).

We followed a consistent process for assignment of measles cases as imported versus locally acquired. First, we reviewed the state or county health department data to see if information was provided on how many annual county-level measles cases were imported or locally acquired. If such data were unavailable, we assigned county-level annual cases into internationally imported or locally acquired when information about the state-level cases allowed unambiguous assignment. There were two scenarios where this was possible: (i) if all state-level cases were either internationally imported or locally acquired, then all county-level cases must also be either internationally imported or locally acquired, and (ii) if all cases in the state occurred in one county, then the state-level apportioning must also be applicable to the county. If there were any differences between the number of international importations reported by CDC and the state health department, we did not assign importations for the state unless condition (i) was satisfied. Over the entire study period, we mapped 52.5% of cases as internationally imported or locally acquired.

To validate our stochastic model of international measles importations, we expanded our dataset to include counties with no measles cases and therefore no internationally imported cases. We knew that if a state-year had no international importations of measles cases, then every county in that year must also have had zero internationally imported measles cases. From 2021-2025, we identified 2,841 county-years as having zero internationally imported measles cases that we added to our validation dataset for our importation model.

*County-level vaccine data*

We obtained data on county-level measles-mumps-rubella (MMR) vaccine coverage, nonmedical exemption prevalence, and medical exemption prevalence in US children from published sources as part of our prior work.[6,7] MMR vaccine coverage was defined as the percentage of children receiving vaccination among all eligible children in the county (details described below).[7] A non-medical exemption is an exemption to the state-based childhood vaccine requirement for personal belief and/or religious reasons.[6] A medical exemption is an exemption to the state-based vaccine requirement due to a medical condition that makes vaccination no longer recommended on clinical grounds.[6] The process used to obtain these data has been described previously.[6,7] In brief, these data were obtained directly from state health or education departments, either through publicly available reports and websites or formal records requests or correspondences. We requested data from 2010-2025 and obtained vaccine coverage, nonmedical exemptions, and medical exemptions for 61.8% of county-years within this time period. Data availability increased over time.

There was some state-level variation in reporting. Most states defined vaccine coverage as the proportion of entering kindergartners receiving two-doses of the measles-mumps-rubella (MMR) vaccine. State-level variation in this definition included: i) defining reported MMR vaccine coverage based on receipt of a single MMR vaccine dose instead of two doses; ii) defining vaccination as being up-to-date for all state required childhood vaccines rather than MMR only; and iii) reporting vaccine data among other age groups, such as K through 5$^{th}$ grade or pre-K through 12$^{th}$ grade. For exemption data, some states reported the number of schoolchildren with exemptions, which we used to calculate a prevalence by dividing by the county's school enrollment as previously described [6,7]. Some states also reported nonmedical and medical exemption prevalences jointly as a total exemption prevalence. Exemption data were sometimes unavailable at a county-level due to privacy reasons from small sample sizes. Non-medical exemptions were issued by states for different reasons such as religious or personal belief reasons but were combined for the analysis if reported separately. Some states only started reporting data in recent years, and some states did not provide recent data.

We performed data quality checks and explored outliers in these data. The nonmedical exemption and medical exemption prevalence data underwent a data review as described previously.[6] In brief, county-years with unexpectedly high nonmedical exemption and medical exemption prevalences in comparison to their MMR vaccine coverage were reviewed, and extreme outliers were removed (<10 county-year observations).[6] We applied a corresponding set of checks to the MMR vaccine coverage data for all counties. We explored outliers in the data defined as <40% coverage and >15 percentage point year-to-year changes in coverage if coverage was <75%. We reviewed the flagged county-years (19 observations), and county-years with exactly zero coverage along with those where the reported coverage could not be reconciled were removed (7 observations).

We calculated alternative measures of vaccination coverage for sensitivity analyses. The primary vaccine data used in our analysis were reported county-level MMR vaccine coverage, nonmedical exemption prevalence, and medical exemption prevalence for the year relevant to the measles outbreak being modeled. We alternatively calculated the population-weighted average vaccine coverage, nonmedical exemption prevalence, and medical exemption prevalence over the last five years in the county to test how time-averaged vaccine data that capture vaccine-derived immunity in children over a broader age group may relate to outbreak size. If five years of data were not available in that county, we used as many years of data as were available. Nevertheless, vaccination coverages between adjacent years and across age groups within the same county are correlated, so the coverage in one year may be an accurate indicator of vaccine-derived immunity among children of multiple age groups. Similarly, immunity among older adults is likely consistent across states and counties given less variation in vaccine uptake in older age groups and since most older adults have infection-acquired immunity.[8]

*Assembling a county-level dataset of vaccination and measles outbreaks*

We joined county-level measles case data with county-level vaccine data to assess the relationship between MMR vaccine coverage and outbreak size. Since vaccine data were gathered among entering schoolchildren generally at the start of the academic year (usually August-September), our inferential analyses sought to relate vaccine data in the year prior to measles cases in the subsequent year (i.e., vaccine coverage in the 2023-2024 academic year would predict outbreaks in 2024). For the few measles cases that were reported at the level of larger spatial units than counties (ex., health districts) or across multiple counties, we aggregated vaccine data by taking the population-weighted average across constituent counties.

We made additional decisions on the use of some county-years with incomplete data. In our inferential analyses to characterize determinants of outbreak size, we dropped county-years where any of vaccine coverage, nonmedical exemption prevalence, and medical exemption prevalence were missing (19.5% of cases/26.6% of county-years) to ensure that we could assess the impact of each of these covariates on measles risk. In a supplementary analysis, we imputed missing vaccine data from data in the same county from neighboring years or state-level data for the year corresponding to the outbreak, reducing dropped cases to 4.7% of cases/8.7% of county-years with confirmation that our inferential results hold over this imputed dataset shown in Figure S7. For the remaining missing locations, we did not have vaccine data from any source that fit our inclusion criteria, and we dropped five county-year equivalents where the cases were not resolved to the level of the county but rather a region because imputation would have required mixing data across sources for these locations.

Because the inferential analyses found that nonmedical and medical exemption prevalence were statistically nonsignificant predictors of outbreak size, when projecting measles risk nationwide, we still made projections in states that reported joint nonmedical and medical exemption prevalences using the total exemption prevalence as the nonmedical exemption prevalence and zero for the medical exemption prevalence since nonmedical exemption prevalence is generally much larger. We used the last available record of vaccine coverage, nonmedical exemption prevalence, and medical exemption prevalence in the county for risk projections, and for counties that never reported data, we used state-wide vaccine data (MMR vaccine coverage or all vaccine coverage, nonmedical exemptions, and medical exemptions) from CDC's SchoolVax viewer.[9]

*County-level sociodemographic covariates*

We selected a list of county-level sociodemographic variables that are plausibly related to measles transmission risk based on prior literature and expert judgement.[10,11] We obtained data from three sources: the US Census's five-year American Community Survey,[12] the US Census 2020 Gazetteer,[13] and the Research Triangle Institute's 2019 synthetic populations of the US.[14] From the US Census, we obtained the population size, fraction of the population under the age of five, fraction of the population that was school-aged, fraction of school-enrolled children attending public school, fraction of the population that is white, fraction of the population above 25 with a graduate degree, and the median income for the year of the outbreak. From the US Census 2020 Gazetteer, we obtained county land sizes to calculate the population density. From Research Triangle Institute's US synthetic populations (which are also reliant on data from the American Community Survey), we obtained the average household size and standard deviation in household sizes across the county.

We performed additional processing on these data. We log transformed the median income and conducted z-score scaling of all variables to accelerate model calibration and convergence. Like with the vaccine data, we calculated population-weighted averages of covariates across constituent counties when the measles case data were reported across multiple counties. For future projections of risk or rare instances where data were not available for the year of an outbreak, we used data from the last available year in that county.

**Additional information on modeling methods**

*Descriptive statistical analyses*

All $p$ values (reported for Spearman correlations and regressions between within-county MMR vaccine variability and multi-county outbreaks) were calculated as permutation $p$ values with a Phipson-Smyth correction[15] where the null distribution was obtained by generating data with equivalent spatial autocorrelation using Moran Spectral Randomization (MSR).[16] This ensured that $p$ values were "spatially aware", meaning that the conclusion of statistical significance accounts for the spatial structure of the data and does not merely reflect spatial clustering. By accounting for unobserved spatial structure in the data, this approach limits Type I errors and is standard when the data have an underlying spatial structure. MSR $p$ values were obtained by generating null data that had the same Moran spectral autocorrelation as the observed data and calculating the empirical $p$ value for observing the patterns seen in our data. We obtained 99,999 random samples for comparison.

*Conditional autoregressive effect at the county-level*

Our main hierarchical Bayesian Poisson regression model included a conditional autoregressive (CAR) effect at the county-level. A CAR effect is a form of a spatial random effect that allows for correlations between county-level random effects based on a global correlation value. The CAR model used in this study was implemented as a multivariate normal distribution (MVN) where the precision matrix was a function of correlation term and adjacencies between counties:[17]

$$CAR \sim s * \text{MVN}(0, D - \rho * A)$$

where $s$ was the standard deviation of the CAR random effect, $\rho$ was the parameter controlling the strength of the spatial dependence between counties, $A$ was the county adjacency matrix, and $D$ was the row-wise sum of $A$ which indicates how many neighbors each county has. County-level adjacency data was obtained from the US Census Gazetteer. Counties that had no neighbors were assigned an uncorrelated, county-level random effect from a normal distribution centered at zero and with a fit standard deviation.

*Time-varying estimates of the relationship of vaccine coverage and outbreak size*

To evaluate how the relationship between MMR vaccine coverage and outbreak size has changed continuously by year, we expanded our main Bayesian Poisson regression model to allow the coefficients for vaccine measures to vary over time via a Gaussian random walk. Because the coefficients for the vaccine data (i.e., MMR vaccine coverage, nonmedical exemption prevalence, and medical exemption prevalence) already varied by state via partial pooling to account for differences in state-level reporting, we made the nationwide (i.e., pooled) coefficient for the vaccine measures vary over time via a Gaussian random walk.

We tested various approaches for modeling the relationship between control covariates and outbreak size in this framework. Our analyses fitting the main Bayesian Poisson regression models independently to 2011-2020 and 2021-2025 suggested that the relationship between control covariates and outbreak size also changed across time periods. As such, our main analysis allowed these variables to vary over time and by state (the most general and akin to how the vaccine data were treated). We sensitivity tested an alternative parameterization to ensure that the extracted relationship between MMR vaccine coverage and outbreak size was robust to this modeling choice where these coefficients could vary over time only. When analyzing the output from this model, we denoised the random walk trend by taking the rolling average over every three time points to facilitate visualization.

In our primary analysis, we present the nationwide coefficient over time for vaccine coverage on outbreak size from the parameterization where control covariates could vary over time and by state using all data averaged with an analysis that removed one clustered outbreak that was an outlier (Hennepin County, Minnesota, 2017). In particular, we conducted sensitivity analyses where we removed outbreaks to evaluate their impact on the time-varying trend. Results from this analysis are shown in Figure S2e-f. We used a similar approach to understand the outbreaks that were most contributing to changes in the relationship between outbreak size and vaccine coverage in 2025.

In this analysis, we assumed the spatial effects stayed constant over time. However, we conducted a supplementary sensitivity analysis where we allowed the CAR random effects to vary independently by year. This allowed the model to include spatial relationships that were time-varying. In particular, we allowed each set of adjacent two years (i.e., 2021-2022, 2023-2024) to have its own independent CAR effect, so a county observed over multiple years could have different spatial relationships at each time point. Our primary goal from this analysis was to assess whether and how the spatial relationships between counties change over time. Results from this analysis are presented in Figure S11e.

*Bayesian calibration of the statistical model*

We used weakly informative or regularizing priors for all parameters. Following the recommendations of Gelman et al., (1995),[18] our hierarchical Bayesian models were implemented using a non-centered parameterization. Applied to our model structure, the state-level coefficients were obtained by taking the nationwide coefficients and adding a state-specific offset, which is calculated by multiplying the state-to-state deviation in the parameter by a state-specific random variate drawn from a standard normal distribution (these are the "standardized" priors in the table below). We used the same approach for modeling the Gaussian random walk of time-varying coefficients: the change between two years was calculated by taking the value in the preceding year and adding the year-to-year temporal drift, which was calculated by taking the year-to-year deviation multiplied by a year-specific random variate drawn from a standard normal distribution. These distributions, along with priors for all parameters, are detailed below in Table S4. Because the data were z-score scaled to accelerate model convergence and enhance comparability between coefficients of different variables, priors were interpreted in z-score space and prior to applying the exponential link function relating linear combinations to Poisson random variates.

We used the No U Turn Sampler Markov chain Monte Carlo algorithm for model calibration. The model was calibrated using four chains run for 4000 iterations each with the first 2000 iterations were discarded as burn-in, yielding 8000 total posterior draws. Chains were fully independent, and for our inferential analyses, each chain was initialized at the median of 15 random samples from each prior, with the exception of one supplementary analysis (rerunning the primary hierarchical Bayesian Poisson model with an additional covariate for previous cases in the county) where we initialized the model at the median of 50 random samples from each prior distribution to speed up chain mixing. In our validation analyses, because we were producing estimates for every US county sometimes with limited training data, we initialized the model at the median of up to 100 random samples from each prior if doing so accelerated convergence. For all inferential analyses, convergence was assessed by requiring a split Gelman-Rubin statistic[19] of <1.01, the absence of positive divergent transitions, an effective sample size of at least 2,000 for each interpreted parameter. For validation analyses, the primary indicator of convergence was the absence of divergent transitions, and larger split Gelman-Rubin statistics were permitted because the goal of these analyses was not interpretation of fit parameters but prediction.

To ensure that all validation models (described in depth below) converged to enable their evaluation, we used modified priors for two parameters (see Table S4 below). We conducted sensitivity analyses where we refit the best-performing model from our validation analyses with the same priors as used in our main analysis, and we found no qualitative difference in our findings (Table S2). We also conducted sensitivity analyses where we used these priors for our inferential analyses, and we found no qualitative differences in our findings.

**Table S4: Bayesian priors for all model parameters.**

| Model | Description | Prior Distribution |
|---|---|---|
| Main + Time-varying | Baseline incidence rate | Inferential: Normal(0, 10.0)<br>Primary validation: Normal(0, 1.0)<br>Sensitivity validation: Normal(0, 10.0) |
| Main | Coefficients for control variables (demographics, year fixed effects, state fixed effects) | Normal(0, 1.0) |
| Main + Time-varying | Mean coefficient for vaccine measures across states (i.e., nationwide; also applied to control covariates in the time-varying model under the variant that control variables vary by time and state) | Inferential: Normal(0, 10.0)<br>Primary validation: Normal(0, 1.0)<br>Sensitivity validation: Normal(0, 10.0) |
| Main + Time-varying | Standard deviation of state-level variation from mean/nationwide vaccine measures | HalfNormal(0.25) |
| Main + Time-varying | Standardized state-specific offsets for the vaccine measures | Normal(0, 1.0) |
| Main + Time-varying | CAR model spatial autocorrelation | 2*Beta(2.0, 2.0) – 1 |
| Main + Time-varying | Standard deviation of spatially structured (conditional autoregressive) random effects | HalfNormal(0.25) |
| Main + Time-varying | Standard deviation of spatially unstructured random effects | HalfNormal(0.25) |
| Main + Time-varying | Standardized spatially unstructured random effects | Normal(0, 1.0) |
| Time-varying | Standard deviation of year-to-year temporal drift in random walk for vaccine measure coefficients | HalfNormal(0.25) |
| Time-varying | Standardized year-to-year offsets in vaccine measure coefficients | Normal(0, 1.0) |
| Time-varying | Standard deviation of state-to-state variations for covariates that vary by state only | HalfNormal(0.25) |
| Time-varying | Standardized state-specific offsets for covariates that vary by state only | Normal(0, 1.0) |
| Time-varying | Coefficients for covariates that vary by time only | Normal(0, 10.0) |
| Time-varying | Standard deviations of temporal drift in random walk for covariates that vary by time only | HalfNormal(0.25) |
| Time-varying | Standardized year-to-year temporal drift offsets in random walk for covariates that vary by time only | Normal(0, 1.0) |

*Relationship between within-county variability in MMR vaccine coverage and multi-county outbreaks*

We obtained school district-level MMR vaccine coverage data to assess the relationship between variability in within-county MMR vaccine coverage and clustering between measles cases in neighboring counties. Variability in within-county MMR vaccine coverage was defined as the standard deviation of school-level MMR vaccine coverage for all schools reporting data within a county for a given year. These data were obtained from a previously published source,[20] and we conducted quality checks on them to ensure they were comparable to our county-level data. In particular, we required that the difference between the county-level vaccine data and unweighted average vaccine coverage across all schools within a county was no greater than 10 percentage points. This removed three county-years. Differences between the two quantities could be from missing data or substantive variation in the population size of each school that required school enrollment data to take the weighted average of school-level data to obtain county-level data.

There was similar variation in the reporting of school-level MMR vaccine coverage by state as there was for the county-level MMR vaccine coverage: in brief, some schools reported MMR vaccine coverage as one dose coverage rather than two dose coverage while some schools only reported coverage for all state-required vaccines, and there was variation in whether the reported vaccine coverage was among entering kindergartners or a different age group. In totality, we obtained data for 126 county-years with measles cases across 2024-2025; these county-years represented 40.6% of measles cases during this time period. The data were unevenly distributed across the two years with 104 county-years in 2025.

We assessed the relationship between variability in within-county MMR vaccine coverage and multi-county outbreaks (quantified as the posterior probability of a spatial relationship between neighboring counties). Our primary analysis used the standard deviation of within-county MMR vaccine coverage as the measure of variability and examined whether this quantity predicted multi-county outbreaks for county-years with 10 or more outbreak cases, and we conducted supplementary analyses to validate that the relationship we find in the main analysis exists across outbreak sizes. We omitted small outbreaks in our primary analysis because we hypothesized that any relationship between the two variables would be insignificant for small outbreaks that were of lower public health relevance since who was infected would be largely stochastic. On the other hand, once the outbreak had spread among a more representative portion of individuals, we hypothesized that homogeneous susceptibility across a county (i.e., low within-county variability), which indicated that the outbreak was more widespread throughout the county, would lead to outbreaks spreading across county borders. To reject the null hypothesis, we needed to find that increasing spatial relationships between counties were associated with smaller variability (more homogeneity) in within-county MMR vaccine coverage. We tested this hypothesis using linear regressions relating within-county variability to the spatial correlation between counties where we ensured that our statistical tests were spatially-aware by using Moran Spectral Randomization to estimate $p$ values.

We conducted supplementary analyses where we validated this relationship across all outbreak sizes. In particular, we used interaction regressions where we interacted outbreak size and within-county variability in MMR vaccine coverage. We focused on the interaction term because we expected the relationship to be increasingly evident at large outbreak sizes where the outbreak had infected a more representative proportion of individuals. To reject the null hypothesis, we needed to find coefficients that suggested that increasing spatial relationships between counties and outbreak size were associated with a smaller variability in within-county MMR vaccine coverage.

In our supplementary analyses, we conducted various sensitivity analyses to ensure our results were robust to analysis choices. We tested an alternative definition of variability in within-county MMR vaccine coverage using the coefficient of variation where the mean MMR vaccine coverage across the county was obtained by taking the unweighted average of the school-level data. We evaluated an alternative measure of outbreak size obtained by taking the logarithm of the outbreak size. We also included state-fixed effects and a control variable for the fraction of schools that were public in the county and the mean MMR vaccine coverage in the county (Table S1).

*Mechanistic model of outbreak size*

Our benchmark (base case comparison) mechanistic model estimated annual county-level local measles cases using the final outbreak size of a susceptible-infected-recovered compartmental model. In this framework, the initial susceptible fraction of people who did not have vaccine-derived immunity is given by $s_0 = 1 - v_c * \eta$ where $v_c$ is the vaccine coverage in the county and $\eta$ is the vaccine efficacy (97% for two-dose MMR). $z = s_0 - s_f$ is the fraction of the population that becomes infected during the outbreak, which is given via the following transcendental equation assuming homogeneous mixing that has a nontrivial solution for $R_0 * s_0 > 1$:[21]

$$s_f = s_0 * \exp(-R_0 * z) \qquad (3)$$

For $R_0 * s_0 \leq 1$, we used a stuttering chains process to estimate the expected value of the outbreak size.[22,23] In our primary analysis, we used estimates of $R_0$ from the literature and assigned the parameter a value of 12.[24] A key goal of our validation analyses was ranking of county-level risk, and the mechanistic formulation used here is robust to the choice of $R_0$. To account for state- and year-level differences in reporting and public health response, we estimated locally acquired cases, $C$, from a Poisson regression of $z$ and state- and year-fixed effects in our inferential analyses.

This approach solves for the final outbreak size from one introduction of an infected individual and therefore assumes that all outbreak cases in a county were part of a single transmission chain. However, in practice, multiple importations could lead to multiple transmission chains. We conducted a sensitivity analysis where we modified Equation 1 when using the output from the mechanistic model to instead multiply our estimate of $C$ by the number of importations, so that the interpretation of $C$ was really the number of local cases *per* importation. In our validation framework, this approach performed worse than assuming that all outbreak cases were part of the same transmission chain (Supplementary Data S1).

*Varying approaches to distributing state-level internationally imported measles cases to counties*

We tested two approaches for modeling county-level internationally imported measles cases from state-level internationally imported measles cases. Our approach for modeling internationally imported measles cases consisted of describing state-level importations as a stochastic Poisson-Gamma (negative binomial) process that was distributed to counties based on a measure of population size. This procedure is equivalent to thinning the state-level process into constituent county-level processes. We tested two measures of population size: (i) the county's population as a fraction of the population of all counties with cases in the state or (ii) the logarithm of the county's population as a fraction of the sum of the logarithms of the population of all counties with cases in the state. We assessed both approaches independently in our validation of the stochastic importation model and picked the approach that had the highest area under the receiver operating characteristic curve (AUC) for predicting whether a county would have at least one international importation in the validation dataset.

*Outbreak model validation*

In our validation framework, we trained our outbreak models on varying rolling window of data prior to predicting case counts in the following year. Although our calibrated outbreak models used in our inferential analyses included state- and year-fixed effects, we did not use these effects in validation model training to ensure generalizability beyond the particular states and years present in the training data. Instead, we tested three time horizons for the length of the rolling window of training data provided to the models: starting 2011 (i.e., training the model on data from 2011-2024 to test on 2025 cases), starting 2019, and starting two years prior to the prediction year (i.e., training the model on data from 2022-2023 to test on 2024 cases and training on data from 2023-2024 to test on 2025 cases). We reported performance from all three testing schemes (Supplementary Data S1, S2, and S3). All models were compared against each other, regardless of whether they had the same training window, to assess what combination of covariates, model structure, and training window yielded the highest performance.

*Further details on model selection*

We evaluated the performance of a total of 13 different outbreak models, each of which were trained on the three time horizons described above. The 13 models included one mechanistic final outbreak size model and 12 hierarchical Bayesian Poisson regression models. In particular, we tested 3 sets of model covariates (shown in Figure 2b): (i) just MMR vaccine coverage, (ii) MMR vaccine coverage, nonmedical exemption prevalence, and medical exemption prevalence, and (iii) all covariates including sociodemographic control variables. For each set of covariates, we tested models with and without the spatial conditional autoregressive random effect at the county-level (2 variants), and we tested models with and without state-level partial pooling in the coefficients for the vaccine data (2 variants), leading to a total of 12 statistical models.

We conducted a model selection process across all models that followed best practices.[25] Our approach used what is considered the standard for model selection: out-of-sample validation where models are tested on different data than the data on which they are trained and their performance on the out-of-sample test data is used to select the best performing model. All outbreak models were joined with the importation model to predict total county-level measles cases. We used a variety of measures to conduct model selection. Our first measure was concordance between observed total nationwide cases and model-predicted average nationwide cases and coverage of the 95% credible interval of model-predicted nationwide cases and observed nationwide cases. For models that had equivalent coverage rates, we prioritized models that had smaller error in predicting outbreak cases (quantified via the root mean squared error of county-level cases). For models that had root mean squared errors (RMSEs) of within 50% of the best performing models, we subsequently assessed the best performing model's ability to predict the particular counties with cases by looking at the minimum validation AUC for predicting whether a county would have at least 10 measles cases, including international importations. The best performing model was chosen as the one with the largest minimum (worst-case) AUC. By looking at both performance in predicting nationwide cases and county-level cases, we ensure that the best performing model balanced both to enhance real-world utility. Finally, we conducted visual inspections between the posterior estimates of nationwide measles cases and observed cases (Figure 4a) and predicted county outbreak risk percentiles and observed cases (Figure 4b). In the main text, we present results from the best performing model (Table 1) and the best performing mechanistic model. We found that the best performing model was the statistical model with all covariates, a spatial conditional autoregressive term, state-specific vaccine measure coefficients, and trained since 2019. The best performing mechanistic model was trained since 2011. Many models performed well.

We calculated the positive predictive value (PPV) and negative predictive value (NPV) based on outbreaks occurring anytime over the validation period rather than in a single year. The prevalence of outbreaks of at least 10 cases was 0.5% across 2021-2025. Because outbreaks are stochastic, we calculated the PPV such that if our model predicted a county as being at risk for an outbreak of greater than a threshold number of cases, if the county had a case in any of the subsequent years, that counted as a true positive for all years rather than assigning that as a true positive only in the particular year when that county had an outbreak. Similarly, we calculated the NPV such that if a county was predicted to not be at risk, it had to never have had an outbreak in any subsequent years to be classified as a true negative. Because the sensitivity and specificity are independent of the prevalence of the positive class, no adjustment was made to how they or the AUC were calculated.

*Sensitivity analyses for the empirical impact of removing nonmedical exemptions*

We repeated our primary analysis for estimating the impact of removing nonmedical exemptions on MMR vaccine coverage independently in each state that has implemented such a policy. We also considered an alternative analysis where we analyzed all states jointly but with state-fixed effects to ensure that our estimated relationship was robust to potential variability in vaccine definitions across states. For all analyses, we assessed the change in MMR vaccine coverage in the year that the policy was implemented, one year after the policy was implemented, and two years after the policy was implemented. Our primary analysis used the relationship between nonmedical exemption prevalence prior to the policy and change in MMR vaccine coverage one year after the policy was implemented, though all coefficients were consistent (Table S3).

*Additional information on future vaccine scenarios and policy intervention scenarios*
We considered a total of six plausible future vaccine coverage scenarios, four of which are presented in the main analysis and two of which were supplementary analyses. The two additional scenarios considered were alternatives of worsening declines and strategies for improvement: instead of vaccine declines doubling, we considered them worsening by 50% (i.e., declines multiplied by 1.5), and instead of reaching universal 95% coverage by 2030, we considered a 1 percentage point increase in year over year coverage to 2030. In all scenarios, nonmedical exemption prevalence and medical exemption prevalence were adjusted accordingly as well. Concretely, medical exemption prevalence always changed at the existing rate in that county, and nonmedical exemption prevalence changed to absorb the changes in vaccine coverage so that the sum of all three could never exceed 100%: for instance, when vaccine coverage declines doubled, nonmedical exemption prevalence increases also doubled.

We considered a total of ten potential policy intervention strategies for increasing vaccine coverage in locations that may be at risk of measles outbreaks, five of which are presented in the main analysis and five of which were supplementary analyses. The five supplementary strategies were alternatives of the five strategies presented main analyses: we assessed increasing vaccine coverage by 10 percentage points in the counties in the bottom decile of vaccine coverage (instead of by 5 percentage points), increasing vaccine coverage by 5 or 10 percentage points in the five states with the highest model-predicted measles risk (rather than removing nonmedical exemptions in these states), and increasing vaccine coverage by 5 or 10 percentage points in the top decile of counties with the highest model-predicted measles risk. In all intervention scenarios, only vaccine coverage and nonmedical exemption prevalence changed: medical exemption prevalence stayed constant. In the strategies where nonmedical exemptions were removed, vaccine coverage increased according to the empirical relationship found in states that removed nonmedical exemptions. In the strategies where vaccine coverage was increased by a given percentage point amount, the increase was done such that vaccine coverage could not exceed 100%. In our simulations, we enforced that increasing vaccine coverage could not spuriously increase outbreak size. For the strategy where we increased vaccine coverage in counties with previous outbreaks (3+ outbreak cases), when the outbreak data were only resolved to the level of a county-equivalent (ex., health district), we did not increase vaccine coverage in all constituent counties as such regions could be quite large. Thus, we likely underestimated the impact of this strategy in practice. Nevertheless, these differences were likely quite small as this constraint removed 12 county-equivalents, and without this, the intervention already impacted 187 counties.

*Additional information on modeling method limitations*
Our time-varying model that estimated how the relationship between vaccine coverage and outbreak size has changed continuously over time was implemented as a random walk where the relationship changed each year but was influenced by neighboring years. While we observed that the estimated relationship was sensitive to particular years, we could not disentangle the impact of each outbreak and the relationship in one year on residual impacts in neighboring years. In particular, we observed that the empirical Spearman correlation between outbreak size and MMR vaccine coverage was weaker in counties that were part of the 2018-19 outbreak in New York and New Jersey.[26] Similarly, as we

describe in the main text, we also noticed a slight attenuation in correlation in 2025. While we identify that smaller outbreaks are responsible for this, we are unable to conclusively determine the mechanism.

Our inferential analyses are designed to detect epidemiologic shifts associated with measles re-emergence, but our predictions are trained on the current epidemiology of measles, which may not be applicable as re-emergence continues with a transition to endemicity. Additionally, as measles re-emerges in the US, there will likely also be domestic importations between counties. We did not explicitly model such importations, and the current model is not designed to model endemicity. This results in our future projections likely underestimating cases, and further data are required to continue tracking the epidemiologic shifts associated with measles re-emergence.

Neither our statistical nor benchmark mechanistic models explicitly included a term for age-specific mixing. This was because we lacked data on the age distribution of measles cases, and we were interested in estimating the total number of annual cases rather than the distribution of those cases over time or by age group.

We conducted a sensitivity analysis using imputed vaccine data for county-years where such data was not directly available, but the main source of imputed data was state-level vaccine data. Additionally, there were more county-years with measles cases lacking vaccine data in the earlier time period (2011-2020). Thus, while we find that our results are replicated in the imputed dataset, it is important to note that there may be additional biases present from using state-level data in this analysis.

*Statistical software*

All analyses were done in Python v3.13. All Bayesian models were implemented and fit in Numpyro v0.19.0. Spearman correlations and linear regression coefficients were calculated using Scipy v1.16.2. Poisson regressions were done using Statsmodels v0.14.5. All code including a complete list of packages and versions used is available in the accompanying Github repository.[27]

#### Supplementary Figures

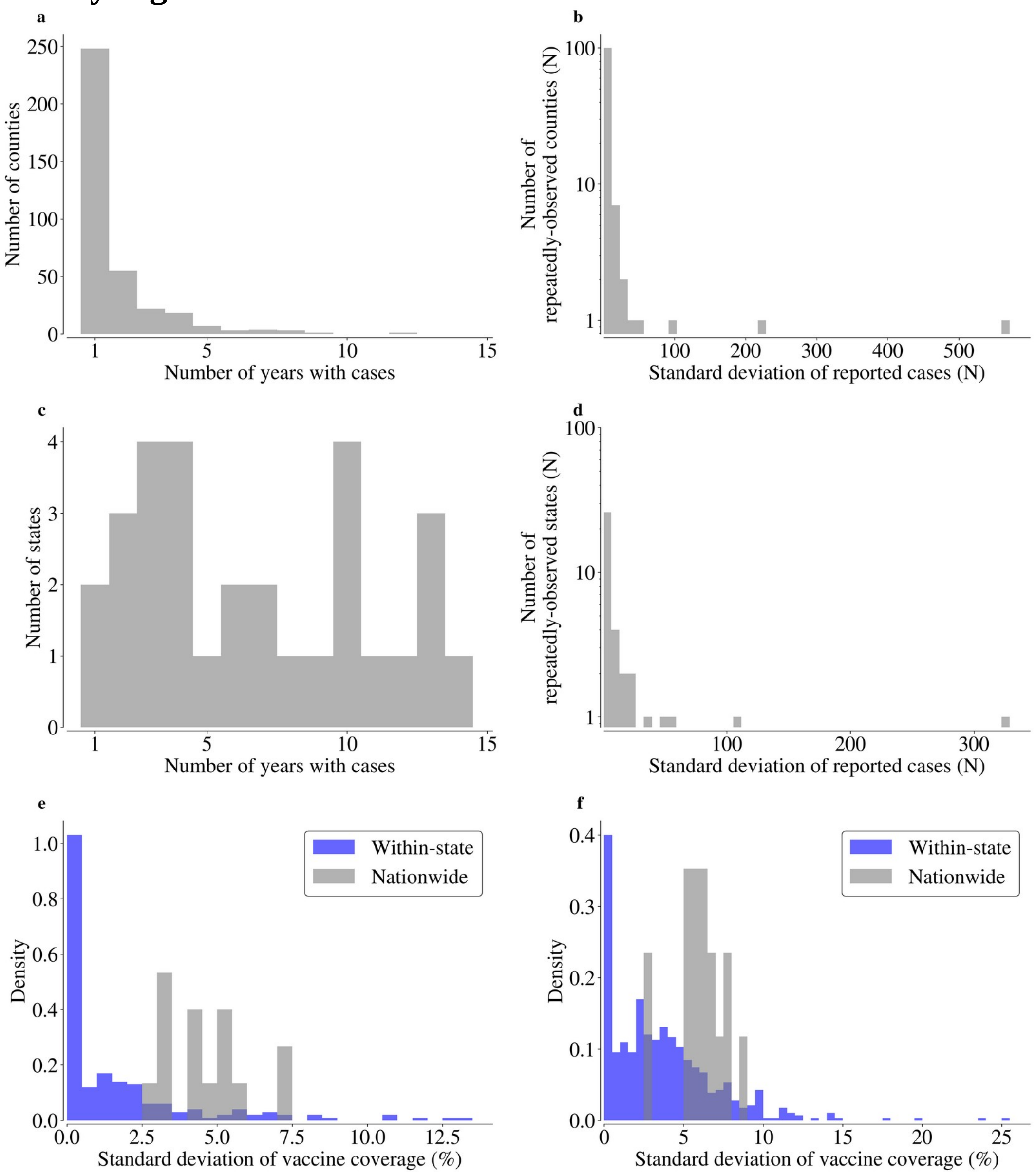


**Figure S1: Variability in measles cases and MMR vaccine coverage by county and state.** (a) Histogram of the number of times a county reports measles cases in the analysis dataset (which requires contemporaneous MMR vaccine coverage and exemption data with the measles case). (b) Standard deviation of outbreak cases (removing international importations) among counties that are observed more than once. (c) As (a) but plotting state-level data instead. Most states are observed at least once (38 out of the 41 states that ever reported measles cases in the dataset). (d) As (b) but for state-level cases. (e) Variability in vaccine coverage within a state compared to nationwide across years. For each state-year, we calculate the standard deviation in vaccine coverage across all counties that had cases in that year; a histogram of this distribution summarizing values from all state-years is shown in blue. We compare this to the standard deviation of vaccine coverage across all counties nationwide that had cases in a given year; a histogram of this distribution summarizing values from all years is shown in grey. (f) As (e) but for all counties with vaccination data, not just those with cases. Taken together, (e-f) indicate that variability in vaccine coverage is much smaller across states than nationwide, motivating state-specific coefficients for the relationship between vaccine coverage and outbreak size.

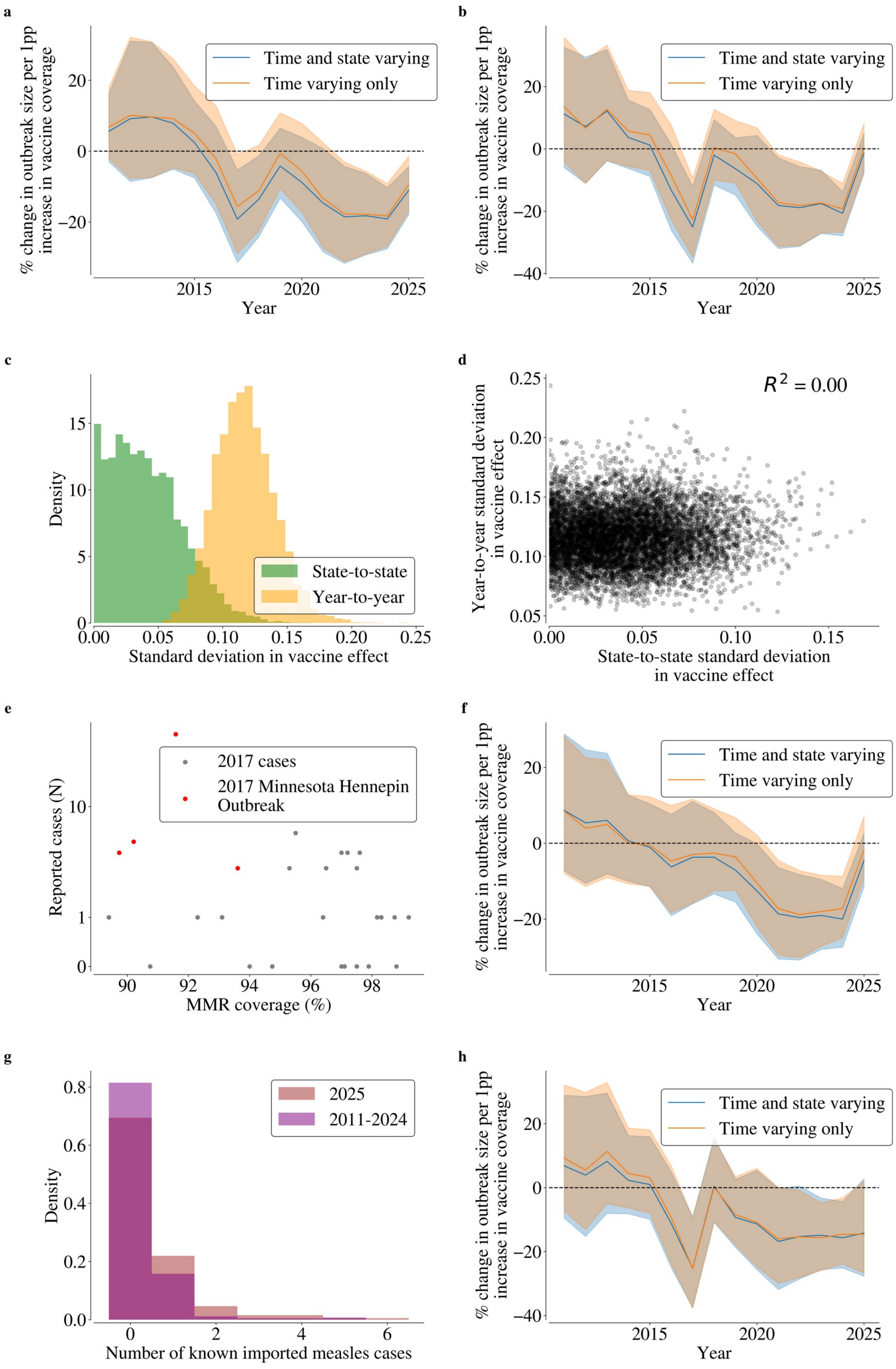
a
% change in outbreak size per 1pp increase in vaccine coverage
Year
Time and state varying
Time varying only
b
% change in outbreak size per 1pp increase in vaccine coverage
Year
Time and state varying
Time varying only
c
Density
Standard deviation in vaccine effect
State-to-state
Year-to-year
d
$R^2 = 0.00$
Year-to-year standard deviation in vaccine effect
State-to-state standard deviation in vaccine effect
e
Reported cases (N)
MMR coverage (%)
2017 cases
2017 Minnesota Hennepin Outbreak
f
% change in outbreak size per 1pp increase in vaccine coverage
Year
Time and state varying
Time varying only
g
Density
Number of known imported measles cases
2025
2011-2024
h
% change in outbreak size per 1pp increase in vaccine coverage
Year
Time and state varying
Time varying only

**Figure S2: Alternative parameterizations of the time-varying relationship between vaccine coverage and outbreak size yield consistent patterns of vaccine coverage becoming an increasingly significant predictor over time.** To ensure the relationship shown in Figure 2a is robust to model specification, we investigated the model posterior and performed sensitivity analyses of different model parameterizations and different data. In both models, the relationship between vaccine coverage, nonmedical, and medical exemption prevalence are allowed to vary by year to appropriately evaluate how the relationship between vaccine indicators and outbreak size has changed continuously over time and over state (as they do in the non time-varying model to account for variability in definitions of vaccine coverage by state). The remaining control covariates (i.e., census-level demographic variables that vary over time) are allowed to be time- and state-varying (blue; same variation as is plotted in 2a) or time-varying only (orange). Consistent trends are observed across both parameterizations. (a) Relationship between MMR vaccine coverage and measles outbreak size over time for all model options, denoised by taking the rolling average over every three time points to facilitate visualization. (b) Same as (a) but without taking the rolling average over the vaccine coverage coefficients, showing a noisier and more stochastic trend. (c) Distribution of the modeled variation of the vaccine coverage coefficient between states (green) and year-to-year (orange) from the primary. We plot these two quantities under the model parameterization in which control covariates may vary over state and time (blue in a) to evaluate whether allowing the vaccine coverage coefficient to vary over time is just absorbing state-level variation in the vaccine coverage coefficient. The standard deviation across time is consistently larger than the standard deviation across states, indicating that changes over time are the dominant force behind deviations in the vaccine coverage relationship, not variations by state. To evaluate whether variation by state may spuriously be modeled as variation over time, (d) shows a pairplot of the posterior samples of the two standard deviations, indicating minimal correlation between the parameters and underscoring that deviations over time and deviations by state are capturing two distinct processes for how the predictive power of vaccine coverage varies. (e) MMR vaccine coverage and outbreak size in 2017, highlighting outlier points part of a clustered outbreak centered in Hennepin County, Minnesota. (f) Removing these points results in a vaccine coverage-outbreak size relationship that shows no sharp discontinuity in 2017 across both parameterizations. (g) Distribution of known imported measles cases in 2025 compared to other years. 2025 had more counties with multiple imports. (h) Removing smaller outbreaks (< 20 outbreak cases) in 2025 that are more influenced by the multiple imports results in no change in modeled relationship between outbreak size and vaccine coverage in 2025 from 2024.

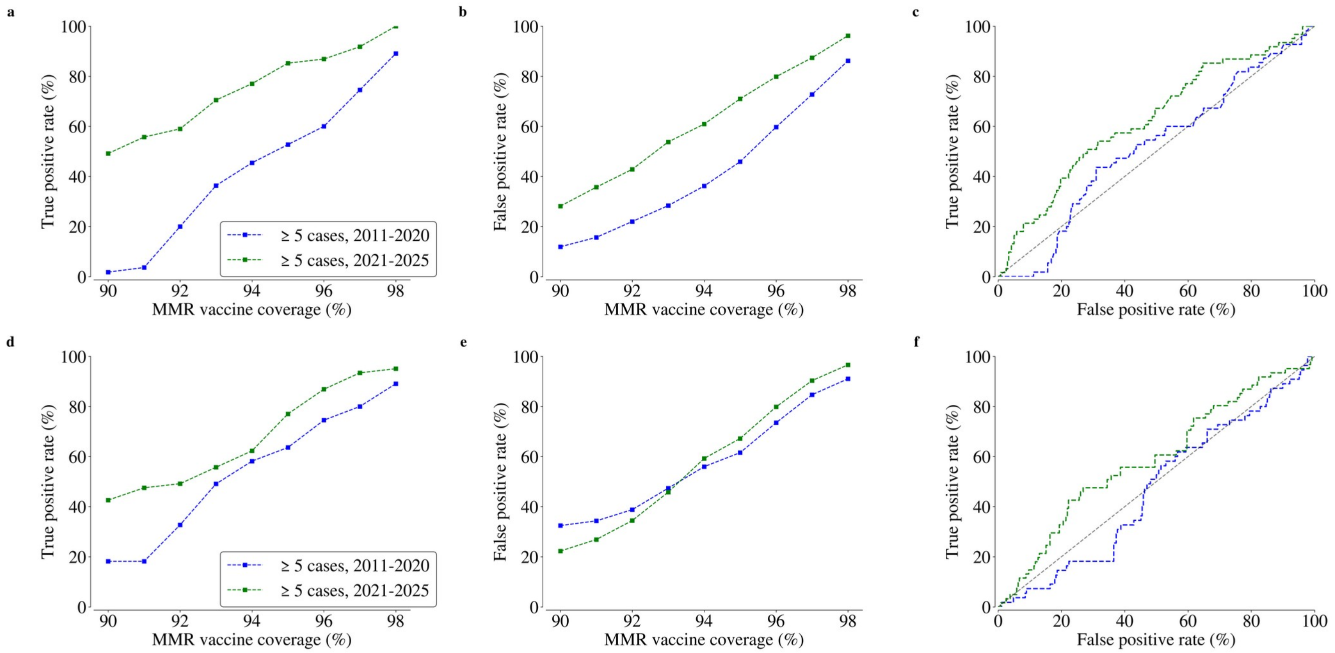


**Figure S3: MMR vaccine coverage alone is a poor classifier of county outbreak risk.** (a) Performance of vaccine coverage thresholds in classifying counties with outbreaks. Points show the identification rate (sensitivity; true positive rate) in classifying measles county-years with an outbreak size of at least 5 cases (i.e., total annual measles cases minus known imports) occurring with MMR vaccine coverage below a given threshold (x-axis) stratified by time period (2011-2020 in blue; 2021-2025 in green). (b) As Figure S3a but plotting false positive rate (100% - specificity) instead of sensitivity/true positive rate. (c) Receiver operating characteristic (ROC) curve from using vaccine coverage as the indicator for whether a county will have at least five outbreak cases. The curve is made by testing different thresholds (like is done in (a) and (b)) and reporting the false positive rate and true positive rate in the corresponding classification of counties. (d-f). Same as a-c except using the population-weighted vaccine coverage over the five years prior to the outbreak as the vaccine measure for classification. If five years of data were not available, as many as are available up to five years were used.

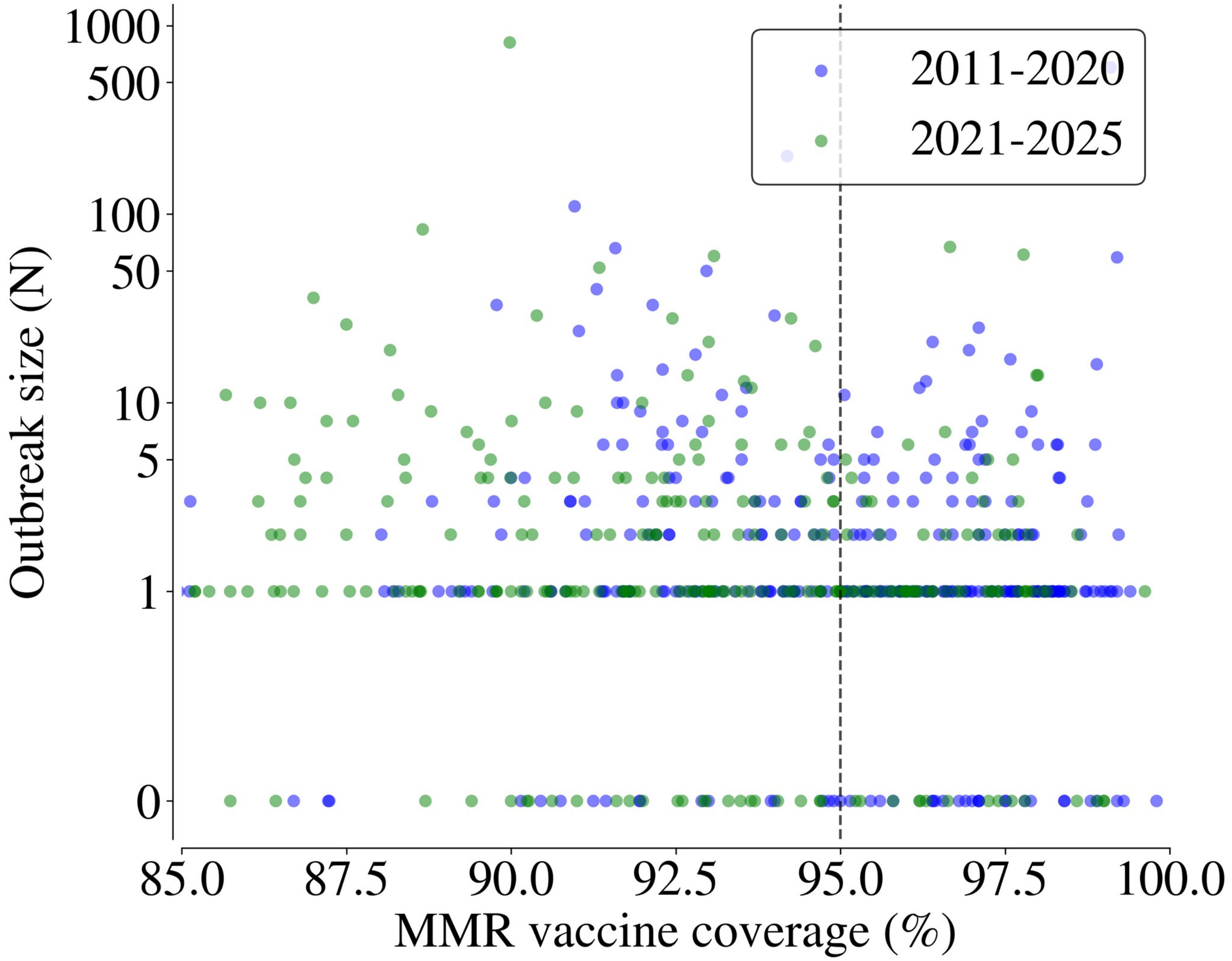


**Figure S4: Outbreak size and vaccine coverage.** Raw data for the violin plots shown in Figure 2b. Outbreak size is the number of total annual measles cases in the county minus number of known international importations, so a value of zero means that there was no local transmission beyond internationally-acquired cases. Points are colored by the time period of the outbreak.

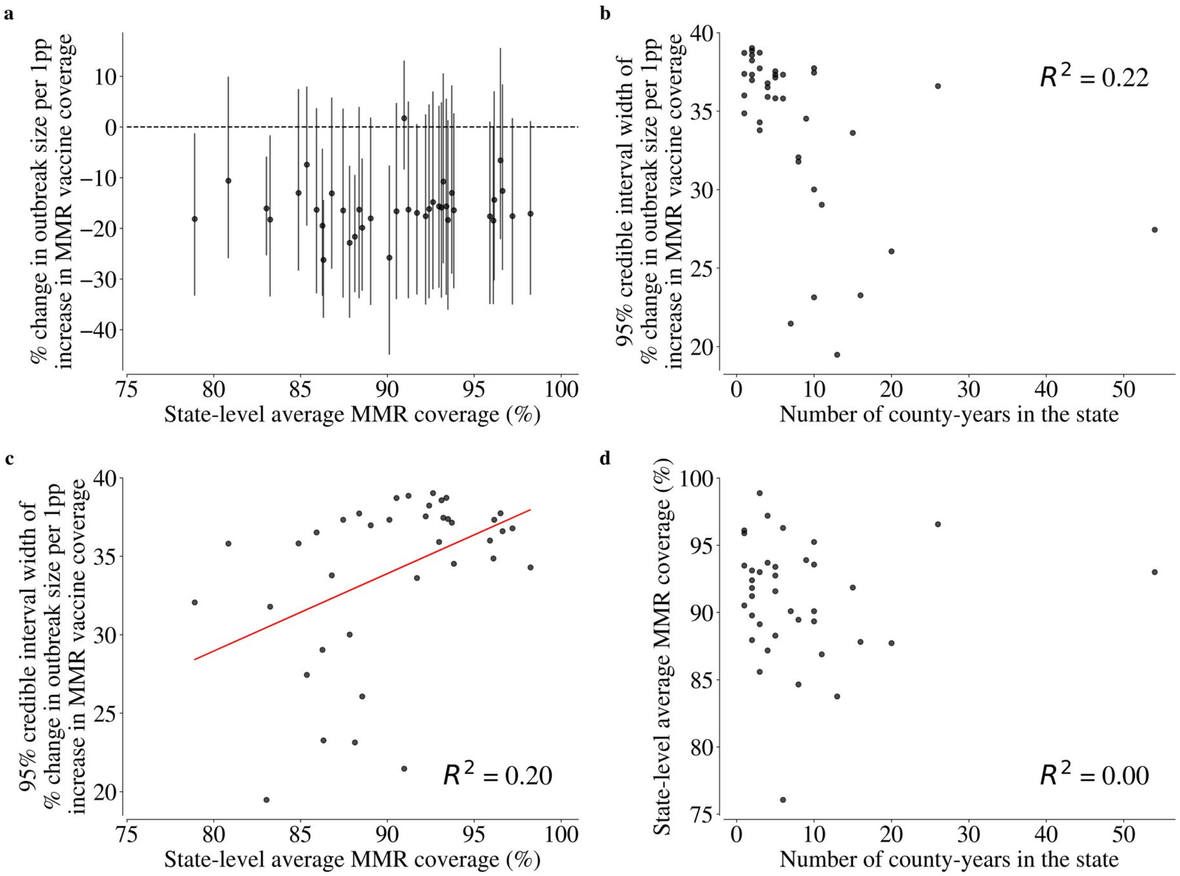


**Figure S5: Hierarchical modeling enables robustly modeling the relationship between vaccine coverage and outbreak size across states.** We investigated how our estimates of the relationship between vaccine coverage and outbreak size vary across states to evaluate the robustness of the relationship and validate whether it follows the patterns expected when using partial pooling in a hierarchical model. (a) Estimated state-level relationship between MMR vaccine coverage and outbreak size as a function of average vaccine coverage across counties that reported cases in that state in 2021-2025. The average vaccine coverage across the state (x-axis) is calculated by weighting the county-level vaccine coverages by the modeled mean case count in that county. Points show the median estimates, and error bars show the 95% credible intervals. There is minimal relationship between the vaccine coverage coefficient and vaccine coverage, indicating that the relationship is robust across states and vaccine coverage. This suggests that the increased association between vaccine coverage and outbreak size shown in recent years is not just because the relationship can only be detected at lower coverages. Credible intervals often contain 0 because there are fewer data points by state than there are nationwide. (b) Relationship between the 95% credible interval width for the vaccine coverage coefficients shown in (a) and the number of county-years present within a state, indicating that increased data enables more precise estimates of the state-specific vaccine coverage coefficient. This is the expected behavior in hierarchical Bayesian models with partial pooling. (c) Relationship between the 95% credible interval width for the vaccine coverage coefficients shown in (a) and average MMR coverage (same x-axis as in (a)). A positive correlation is observed, consistent with greater uncertainty in the impact of vaccine coverage as vaccine coverage is higher and outbreak risk is reduced. (d) To ensure the relationship in (c) is not because of a spurious correlation between average MMR coverage and number of county-years in the state (shown in (b)), we plot the relationship between number of county-years in a state (x-axis in (b)) and average MMR coverage (x-axis in (c)).

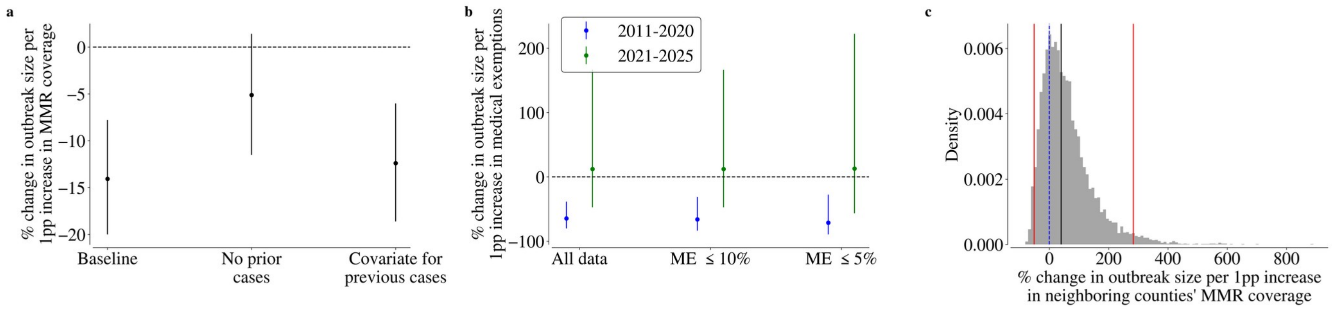


**Figure S6: Sensitivity analyses of the relationship between vaccine coverage and outbreak size.** To ensure the estimated relationships between vaccine coverage, exemptions, and outbreak size were robust, we refit our model under different constraints on the underlying outbreak data. (a) Estimated vaccine coverage coefficient is robust to previous outbreaks in a county. We test different ways of incorporating the impacts of previous outbreaks on our estimated vaccine coverage coefficient from the Bayesian Poisson model with all covariates for 2021-2025. "Baseline" shows exactly what is shown in Figure 2b's yellow bars for reference. "No prior cases" refers to fitting the model to only the first outbreak for each county-year (if duplicated), ensuring that the county had never previously experienced an outbreak over the study period but limiting the dataset. "Covariate for previous cases" refers to fitting the model to all county-years in the time period but including a covariate for the previous cases in that county (even prior to 2021). In all cases, we see that the vaccine coverage coefficient remains negative and is significant when including a covariate for previous outbreak size. While the coefficient borders on significant ($p$ = 0.06) when limiting the dataset to just the first outbreak in a county, doing so removes 36% of county-years. (b) Relationship between medical exemption prevalence and outbreak size is robust to outlier values of medical exemptions. Estimated relationship of medical exemptions on outbreak size from the Bayesian Poisson regression with all covariates fit independently to data from both time periods when limiting to county-years with particular medical exemptions. "All data" shows the same quantities plotted in Figure 3b, "ME ≤10%" refers to only including county-years with medical exemptions less than or equal to 10%, and "ME ≤5%" refers to only including county-years with medical exemptions less than or equal to 5%. Coefficient values remain consistent despite subsetting and removing large values of medical exemption prevalence, suggesting that the protective association between medical exemptions and outbreak size from 2011-2020 and the lack of relationship between medical exemptions and outbreak size from 2021-2025 are not driven by outlier counties. (c) MMR vaccine coverage in neighboring counties does not significantly predict outbreak size. Using the Bayesian Poisson regression model with all covariates, we add the average MMR vaccine coverage in neighboring counties as a control covariate to understand whether the importance of the conditional autoregressive spatial effects is explainable by similarities in neighboring county's vaccine coverage. The large credible interval suggests that vaccine coverage in neighboring counties is alone not predictive of outbreak size in the focal county, though the median impact is negative.

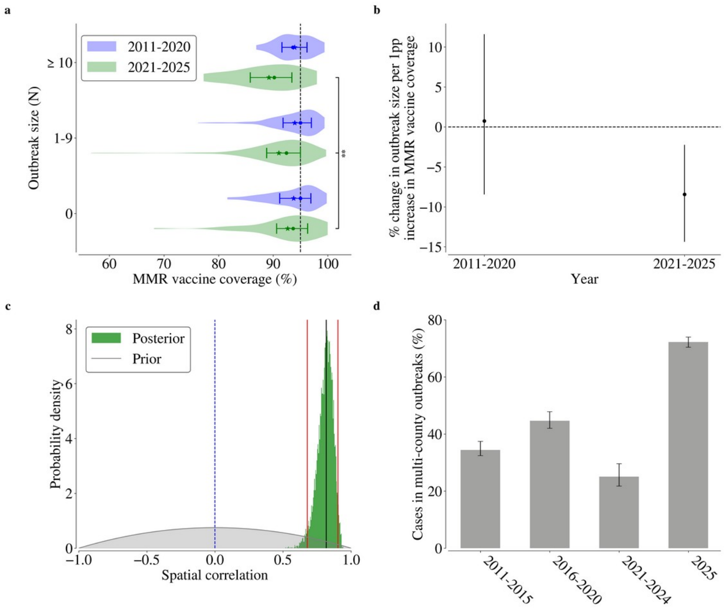


**Figure S7: Inferential analyses suggesting a shift in measles epidemiology in recent years are reproduced when imputing vaccine data in counties where such data is unavailable.** To assess the robustness of our analyses showing a strengthening of the relationship between vaccine coverage and outbreak size and an increase in multi-county spatially related outbreaks in recent years, we repeated our analyses including imputed vaccine data, resulting in reducing county-years with measles cases and unavailable vaccine data from 26.7% to 8.6%. (a) County-level MMR vaccine coverage by measles outbreak size across time periods. This panel is structured as Figure 2b. Points show the median MMR vaccine coverage across observations in each category, stars show the mean, and error bars show the 25$^{th}$ and 75$^{th}$ percentiles. The horizontal dashed line is drawn at 95% vaccine coverage, the targeted critical immunity threshold. The asterisks indicate that vaccine coverage and outbreak size were significantly correlated in the 2021-2025 period (Spearman $\rho$ = -0.16; $p$ = 0.007) but not in the 2011-2020 period (Spearman $\rho$ = -0.03; $p$ = 0.6). (b) Adjusted estimate for relationship between county-level MMR vaccine coverage and measles outbreak size using Poisson regressions with all covariates fit independently to outbreak data from the two time periods. The model includes state- and year-fixed effects and a conditional autoregressive spatial random effect at the county level. Coefficients for MMR coverage, nonmedical exemptions, and medical exemptions can vary by state to account for state-level variation and reporting differences. Points show the posterior median of the estimated nationwide relationship between MMR vaccine coverage and outbreak size, and error bars show the 95% credible interval. (c) Measles cases between neighboring counties were spatially correlated from 2021-2025. Posterior distribution of the conditional autoregressive autocorrelation term from the Bayesian Poisson regression model with all covariates fit to outbreaks from 2021-2025 (green). The grey distribution shows the prior where we assumed there was no spatial autocorrelation. The blue dashed line marks a spatial autocorrelation of 0. The black line shows the median and red lines show the 2.5% to 97.5%. The spatial autocorrelation describes the degree to which the random effects are correlated between neighboring counties. A positive value suggests that when one county has a larger outbreak size than expected, its neighbors do too. Counties may have greater outbreak sizes than expected because of county-county spillover or unobserved spatially-clustered characteristics that are relevant for calibration. (d) Spatial clustering of measles cases over time. This panel is structured as Figure 3c. We calculated the proportion of measles cases in counties that had at least three non-imported cases where a neighboring county also had at least three cases in the same year ("multi-county outbreaks"; y-axis) averaged across years in various time periods (x-axis). Error bars show 95% credible intervals from a beta sampling distribution.

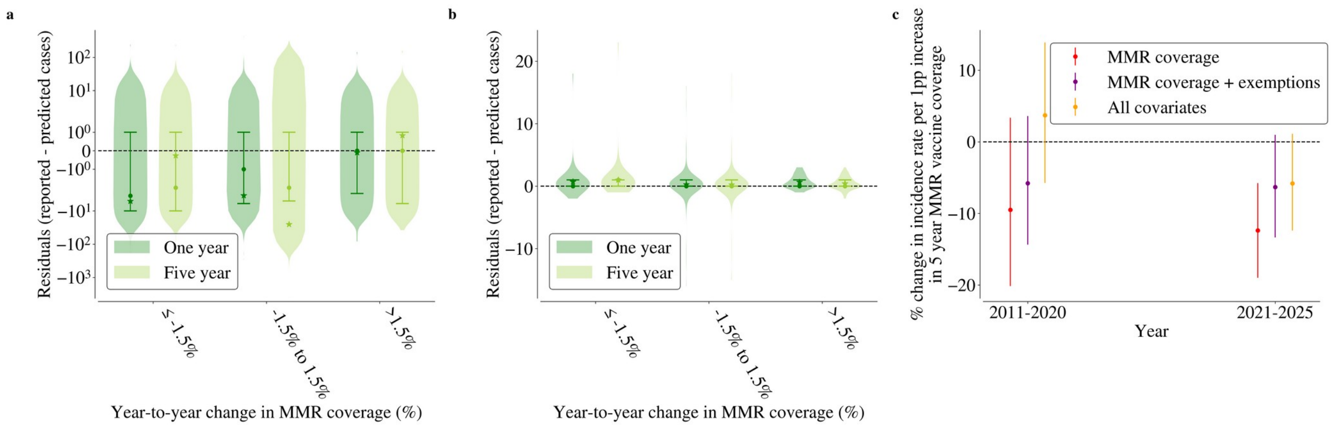


**Figure S8: Outbreak size estimates are robust to alternative measures of vaccine coverage across larger age groups.** In our main analysis, we modeled outbreak size using the vaccine coverage in the year corresponding to the outbreak ("one year"). As a sensitivity analysis, we investigated the calibration performance of our model when using the population-weighted vaccine coverage in the five years prior ("five year"). If five years of vaccine data were not available in the county, we used as many were available. (a) and (b) compare the calibration performance (quantified as residuals of the reported minus predicted cases; y-axis) of the mechanistic (panel a) and Bayesian Poisson model with all covariates (panel b) when using the one year or five year coverages. The Bayesian Poisson model also included five year averages of nonmedical and medical exemption prevalence. We found that calibration performance for each of the two models was robust to different measures of vaccinations, and this was maintained across varying year-to-year changes in vaccine coverage (x-axis). Points show the median of the residual distribution, stars show the mean, and error bars show the 25$^{th}$ to 75$^{th}$ percentiles. However, (c) shows the estimated relationship between the population-weighted five year vaccine coverage and outbreak size from the Bayesian Poisson model. This panel is the same structure as Fig. 2b but with the population-weighted five year prior average as the vaccine measure in the model. Error bars indicate the 95% credible intervals. The vaccine coverage coefficient is smaller and less significant in 2021-2025, suggesting that five year vaccine measures are less sensitive to the changing relationship between vaccine coverage and outbreak size. Nevertheless, a strengthening of the relationship is still observed, underscoring an epidemiologic shift in the explainability of measles outbreaks.

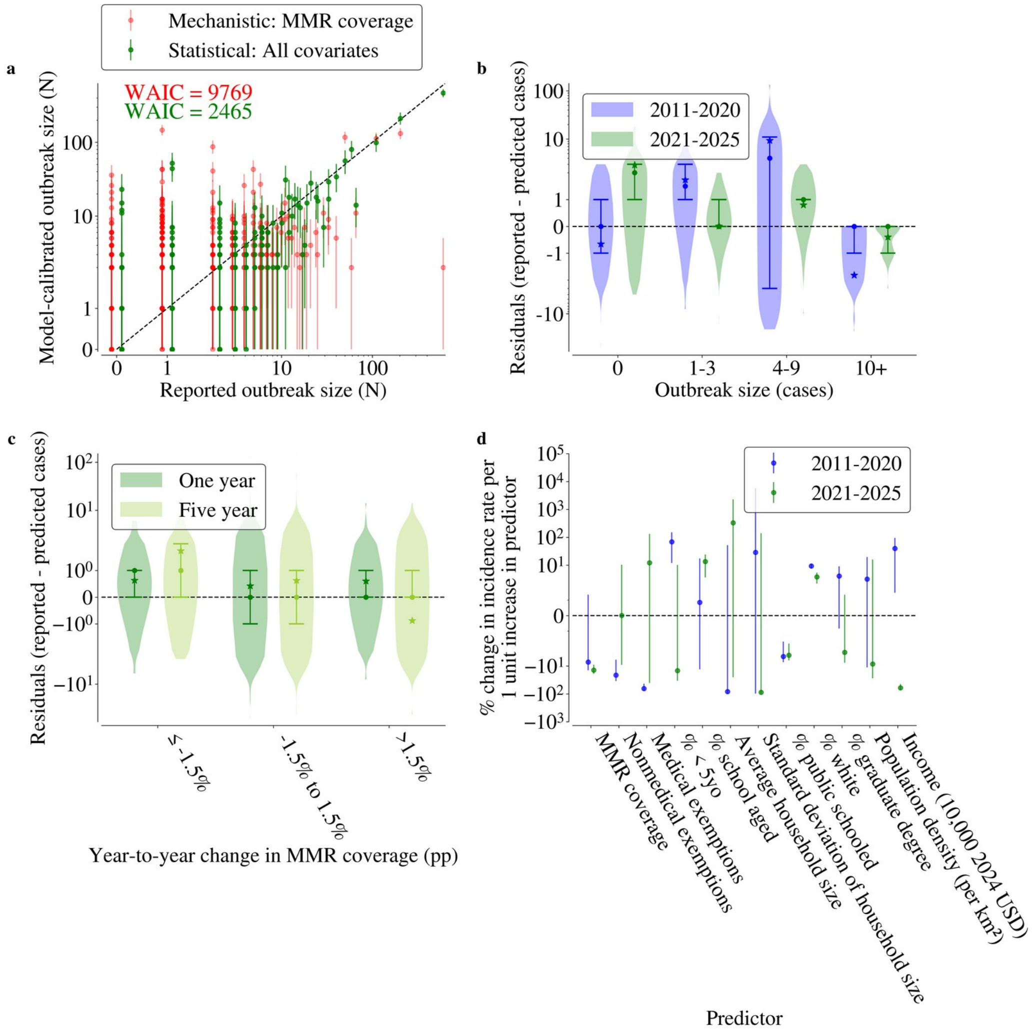


**Figure S9: Calibration performance for 2011-2020.** (a) Calibration model performance predicting measles cases for all county-years reporting cases from 2011-2020. Green points show the posterior median estimates from a hierarchical Bayesian Poisson regression model based on MMR vaccine coverage and sociodemographic covariates, state and year fixed effects, and a conditional auto regressive spatial random effect at the county level. Red points show the median estimate from a final outbreak size mechanistic model based on county-level vaccine coverage. For both models, error bars show the 95% credible intervals. Model performance in a validation framework shown in Table 1 and Figure 4. This panel is structured as Figure 3a but for 2011-2020. (b) Calibration performance is better in 2021-2025 than 2011-2020 for the Bayesian Poisson model. Violin plots showing the distribution of residuals (reported minus median predicted cases; y-axis) as a function of outbreak size (x-axis) and time period (colors) for the Bayesian Poisson regression shown in Figure 3a and S7a. Stars indicate the mean; points indicate the median of the distributions. Error bars show 25$^{th}$ to 75$^{th}$ percentile ranges. (c) Residuals (y-axis) by average year-to-year change in MMR coverage when modeling with vaccine coverage and exemptions in the prior year versus the population-weighted five year prior average for 2011-2020. This panel is structured as Figure S5b but for 2011-2020. Stars indicate the mean; points indicate the median of the distributions. Error bars show 25$^{th}$ to 75$^{th}$ percentile ranges. (d) Estimated relationships between outbreak size, vaccine coverage and exemptions, and all socioeconomic and demographic covariates for the 2021-2025 time period. This panel is structured as Figure 3b but for all covariates, not just those that are significant from 2021-2025.

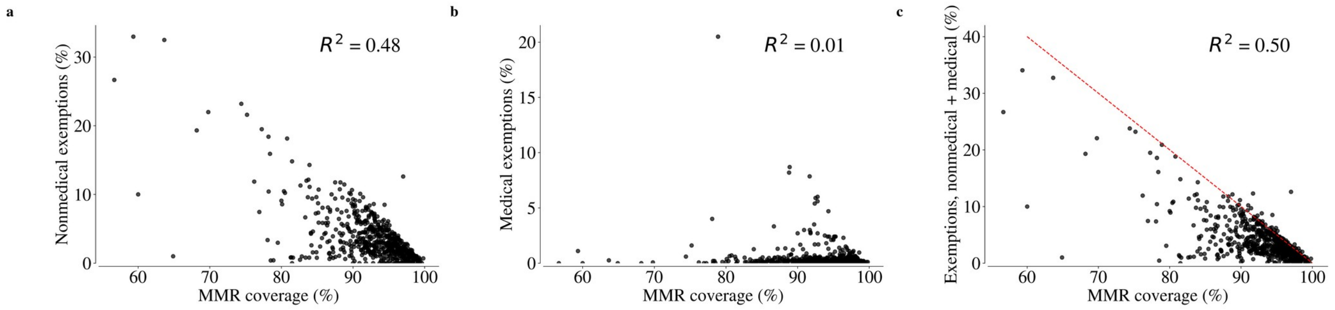


**Figure S10: Vaccine coverage and nonmedical exemptions are correlated while vaccine coverage and medical exemptions are not.** (a) County-level vaccine coverage and nonmedical exemption prevalence for all counties in the analysis dataset from 2021-2025. A strong negative relationship is observed, indicating that increasing MMR coverage is associated with decreasing nonmedical exemptions. This suggests that the lack of significance for the relationship between outbreak size and nonmedical exemptions (Figure 3b) is because nonmedical exemptions do not provide any additional information due to correlation between the two variables. (b) County-level vaccine coverage and medical exemption prevalence for all counties in the analysis dataset from 2021-2025. Compared to (a), no relationship is observed between MMR coverage and medical exemptions. This suggests that medical exemptions provide complementary information to MMR coverage. While most counties have low medical exemptions, some have high values, so we ran sensitivity analyses where the model is re-calibrated on subsets of the data with lower values of medical exemptions (shown in Figure S6c). (c) County-level vaccine coverage and total exemptions (nonmedical plus medical) for all counties in the analysis dataset from 2021-2025. The red line shows the theoretical line where coverages and exemptions sum to 100%. The majority of counties are below the red line, indicating that the vaccination status of some individuals is unknown. Any counties above the red line have coverage plus exemptions that sum to over 100%. This can arise from children who obtained nonmedical exemptions while they were waiting to get vaccinated and their nonmedical exemptions have not yet cleared, or children who obtained nonmedical exemptions for some vaccines other than MMR.

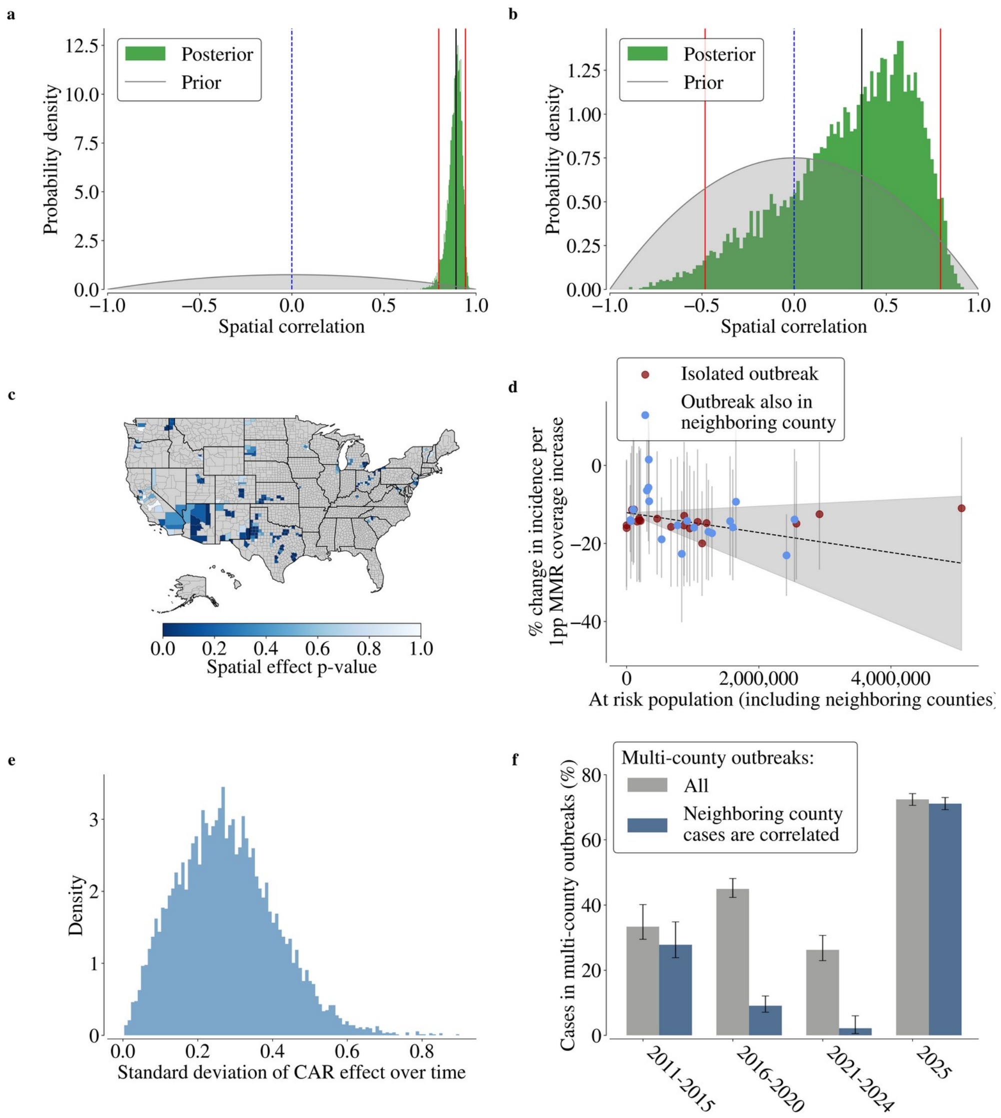


**Figure S11: Measles outbreaks are spatially clustered.** (a) Measles cases between neighboring counties were spatially correlated from 2021-2025. Posterior distribution of the conditional autoregressive autocorrelation term from the Bayesian Poisson regression model with all covariates fit to outbreaks from 2021-2025 (green). The grey distribution shows the prior where we assumed there was no spatial autocorrelation. The blue dashed line marks a spatial autocorrelation of 0. The black line shows the median and red lines show the 2.5% to 97.5%. The spatial autocorrelation describes the degree to which the random effects are correlated between neighboring counties. A positive value suggests that when one county has a larger outbreak size than expected, its neighbors do too. Counties may have greater outbreak sizes than expected because of county-county spillover or unobserved spatially-clustered characteristics that are relevant for calibration. (b) Same as (a) but for 2011-2020. (c) Map of the Bayesian $p$-value of the conditional autoregressive spatial random effects fit for each county from 2021-2025 with at least one measles case. (d) Spatial spillover risk is associated with increasing association between vaccine coverage and outbreak size. The y-axis shows the estimated vaccine coverage coefficient for each state. The x-axis shows the population of the largest set of multi-county outbreaks (contemporaneous and at least 3 cases in neighbor) occurring within that state. Points are colored by whether the state ever had a county that was part of a clustered outbreak. The relationship between the vaccine coverage coefficient and at risk population is significant for states that had outbreaks in neighboring counties (Spearman $\rho$ = -0.4; $p$ = 0.045) but is not significant for states that did not ($p = 1.0$). The dashed line shows a Theil-Sen robust estimator regression for this relationship for states that had a county with an outbreak in a neighboring county with the shaded areas showing 95% confidence intervals. This suggests that vaccine coverage has become a more significant predictor of outbreak size as outbreaks have spread more among counties. (e) Variation in the conditional autoregressive random effect at the county-level over time from the supplementary sensitivity analyses where we allowed the CAR effect to vary over time. While there is some variation, it is moderate, and the majority of posterior samples are at lower values, indicating that spatial relationships have likely not changed over time but just that their strength has. (f) Spatial clustering of measles cases over time. This panel is structured as Figure 3c, but it also shows the percentage of multi-county outbreaks where the CAR effects were significant (blue). We calculated the proportion of measles cases in counties that had at least three non-imported cases where a neighboring county also had at least three cases in the same year ("multi-county outbreaks"; y-axis) across time periods (x-axis). We subset that percentage to include only multi-county outbreaks where at least one county had a significant CAR effect (blue). Error bars show 95% credible intervals from a beta sampling distribution.

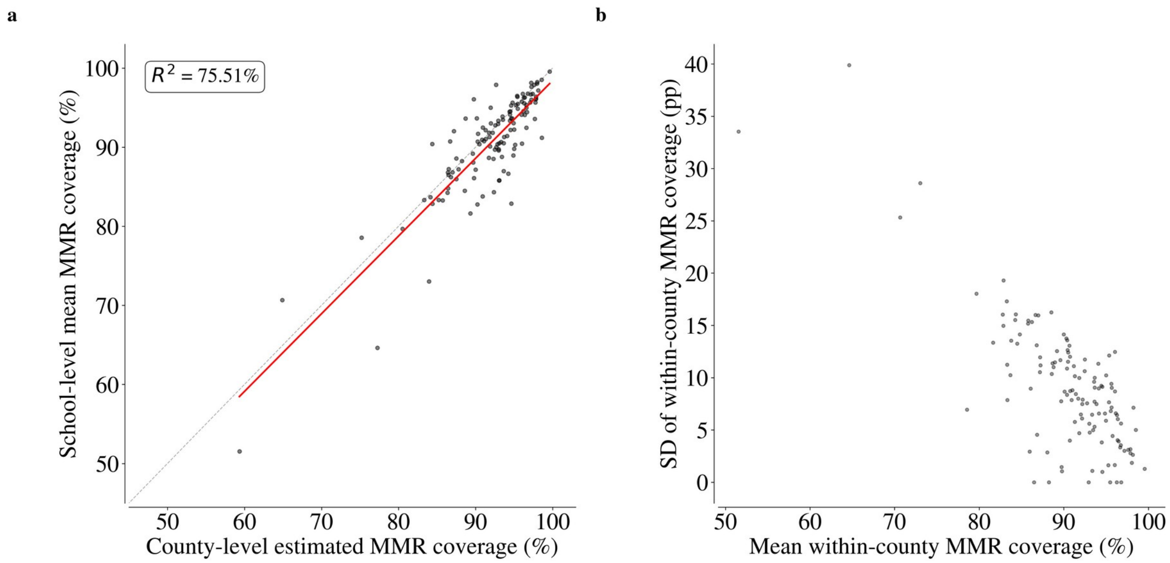


**Figure S12: Descriptive analyses of within-county MMR vaccine coverage and comparisons to county-level data.** (a) County-level vaccine coverage and average school-level vaccine coverage for schools within a county. For available counties with school-level vaccine coverage data ($n$ = 126 across 2024-2025), we compared the population unweighted average MMR vaccine coverage across schools within a county (y-axis) to the county-level MMR vaccine coverage (x-axis). The grey line is the 1:1 line, and the red line is the best fit line with an $R^2$ of 75.51%. (b) Relationship between variability in within-county MMR vaccine coverage and mean within-county MMR vaccine coverage. For counties in our dataset, we calculate the population unweighted average MMR vaccine coverage across schools within a county and relate that to the unweighted standard deviation of MMR vaccine coverage across schools within a county.

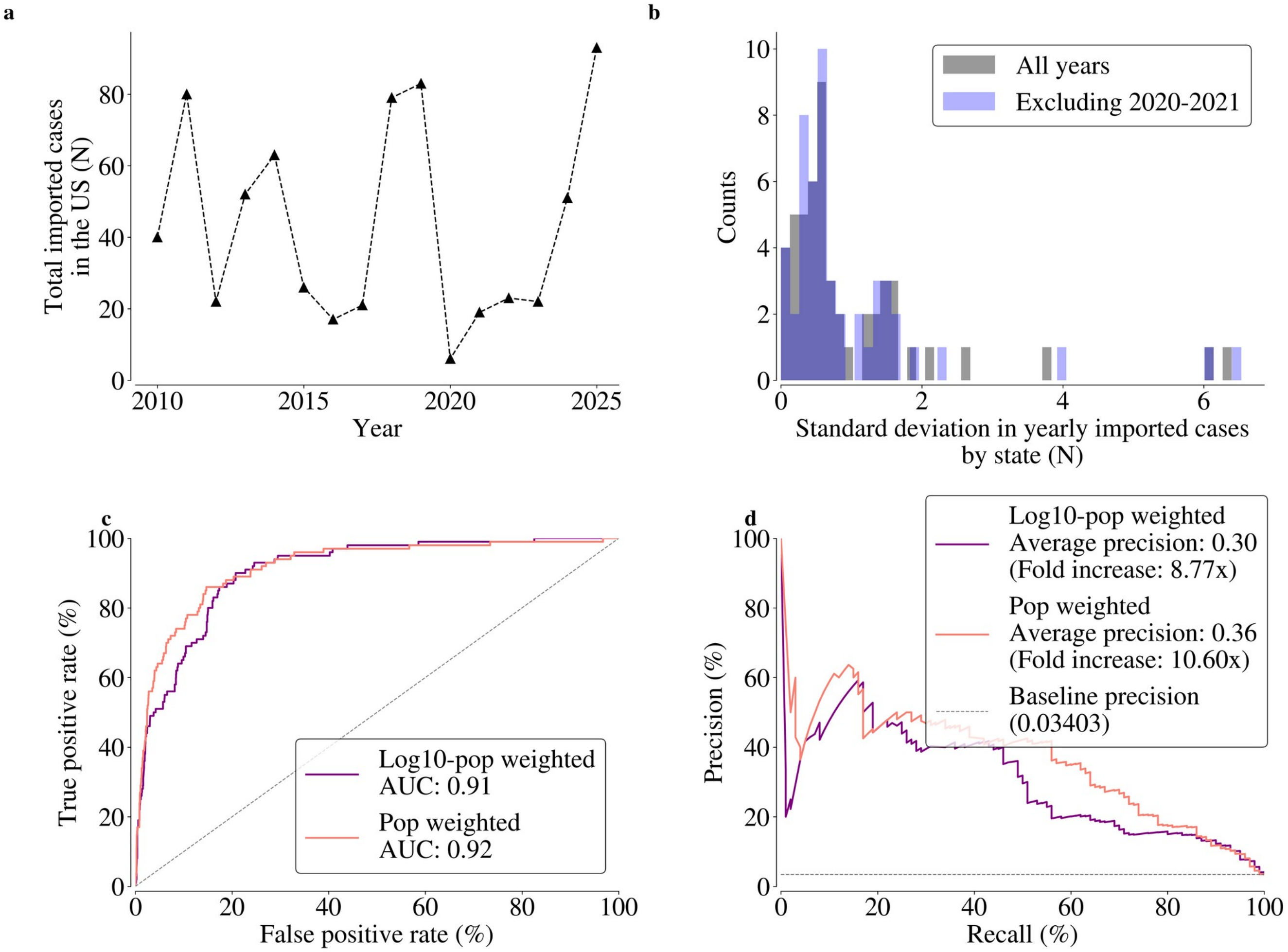


**Figure S13: A stochastic model of measles importations reproduces county-level observed measles importations.** (a) Imported measles cases nationwide by year. Yearly measles imports fluctuate randomly over time, but the range of fluctuations is constrained. (b) States consistently have similar numbers of measles imports over time. For every state, we calculate the standard deviation of imported cases reported over all years (histogram shown in grey) and when excluding 2020-2021 (COVID years when international travel was decreased; histogram shown in blue). Both distributions are clustered at low standard deviations, indicating that states see consistent numbers of measles imports over time. (c) Validation receiver operating characteristic curves for predicting whether a county will have at least one import in 2021-2025 for the stochastic measles model that uses state-level imports and county population calibrated to data from 2011-2020. The model uses a Bayesian estimation process to calibrate state-level Negative Binomial processes for the number of yearly imports and then distributes those processes to the county-level based on either county population (pink) or the logarithm of the county population (purple). (d) Same data and methodology as (c) but showing a precision-recall curve instead of a receiver operating characteristics curve.

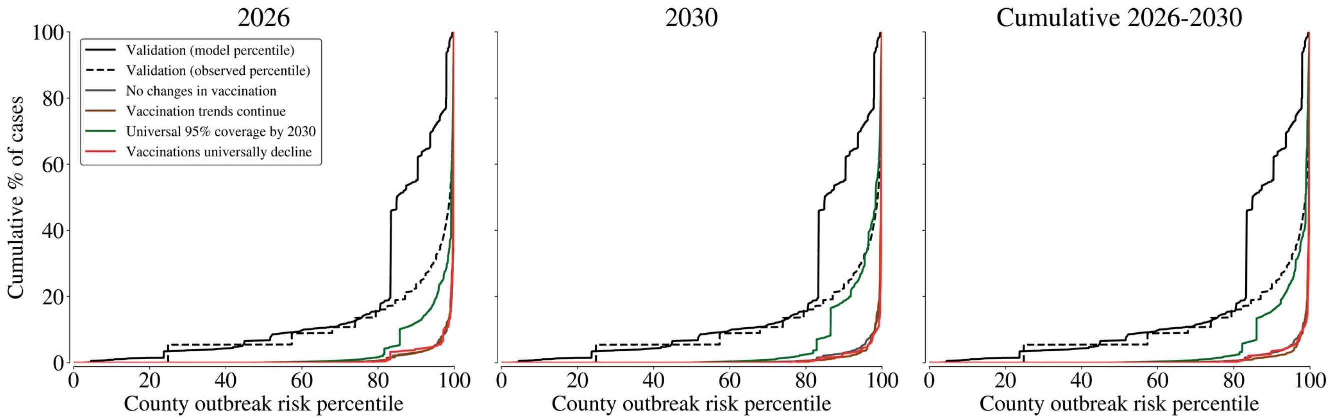


**Figure S14: Validation analysis of measles model reproduces outbreak size distribution and matches county's outbreak size percentile.** All panels show the same validation lines of model performance on withheld data from 2021-2025. The solid black line shows the cumulative distribution function of the model-predicted county-level outbreak sizes while the dashed black line shows the same quantity but using the observed outbreak size percentile to rank-order counties (the y-axis still shows the predicted cases from that county). Concordance between the two black lines shows strong model performance of not only reproducing the observed outbreak size distribution but matching each county's empirical outbreak size percentile. In particular, we show that the top 20% of counties by risk ranking contain 80% of cases. The colored lines show model-projected county-level outbreak size distributions under various vaccine coverage scenarios in 2026, 2030, or aggregating all projected cases cumulatively from 2026-2030. Each scenario shifts the outbreak size distribution modestly, but all scenarios show that risk is spatially localized with the vast majority of cases in a minority (~10%) of counties, underscoring the value of spatially targeted interventions.

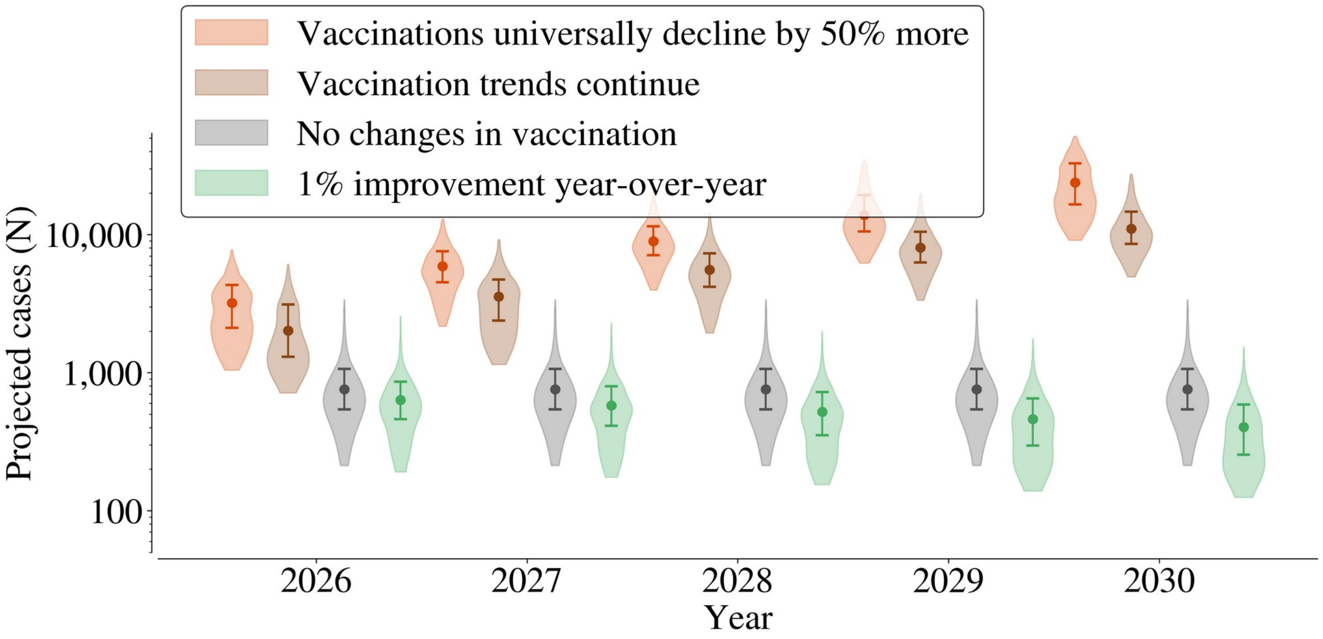


**Figure S15: Future case projections under alternative scenarios of changes in vaccine coverage.** Projected annual nationwide measles cases 2026-2030 under various vaccination scenarios, using a log axis. "Vaccine trends continue" refers to continuation of the trends presented in Figure 1d. "Vaccination universally decline by 50% more" refers to switching counties whose coverage is increasing to instead decline at the national average and increasing the rate of decline in all counties experiencing declines by 50%. "1% improvement year-over-year" refers to all counties experiencing a 1% increase in vaccine coverage until their coverage hits 100%.

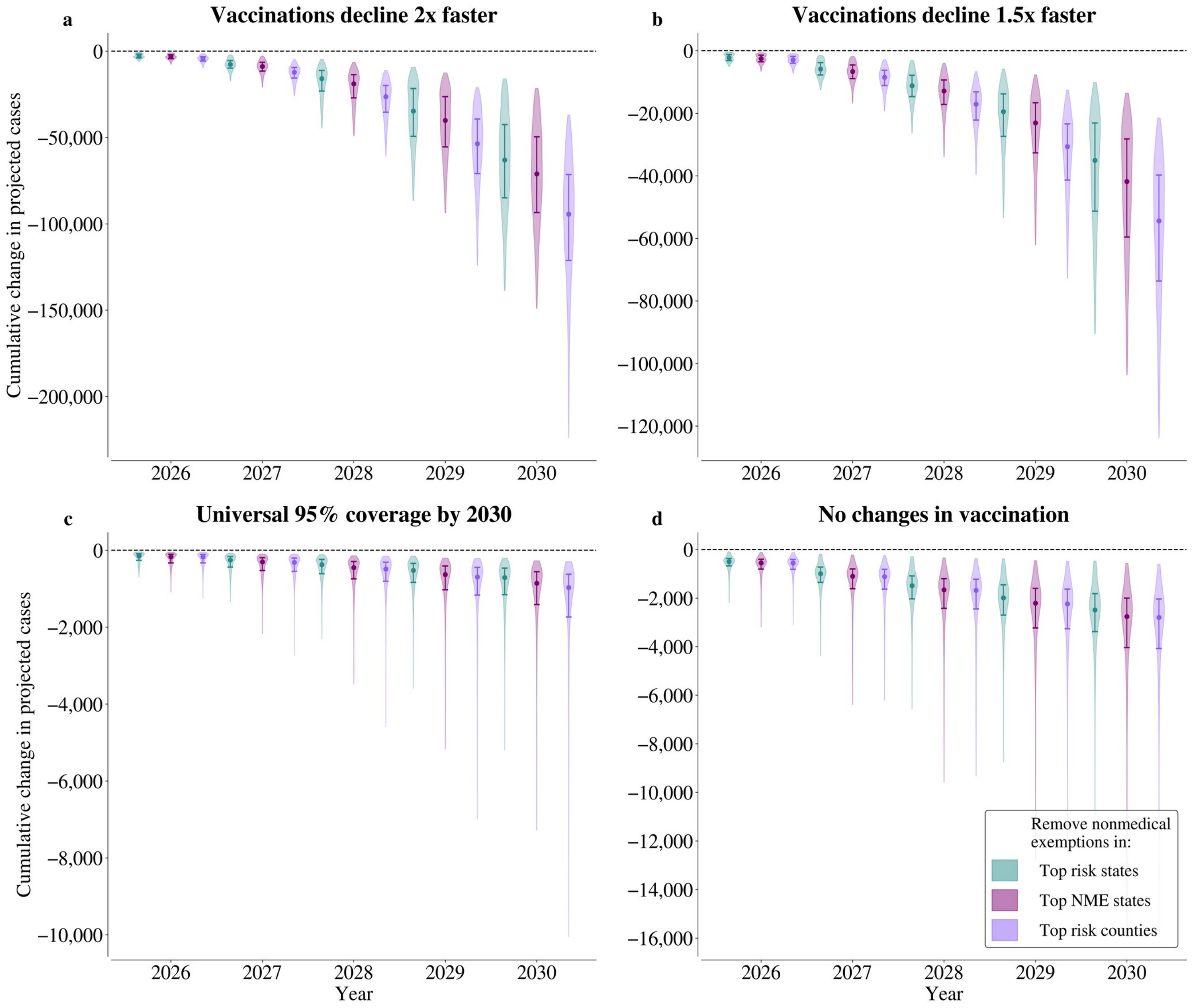


**Figure S16: Sensitivity analyses of impact of NME policies under different future MMR vaccine scenarios.** Panels show the projected impact of various policy strategies to increase vaccine coverage over time. We show the results from three such strategies: removing nonmedical exemptions in the five states with the highest nonmedical exemption prevalence, removing nonmedical exemptions in the five states with the highest model-predicted measles cases, and removing nonmedical exemptions in the top 10% of counties with the highest predicted outbreak sizes. Violins show the 95% posterior range of cumulative change in cases over time, points indicate the median, and error bars show the middle 50% posterior range. Each panel shows the projected impacts under different scenarios of change in vaccine coverage. (a) Vaccination declines universally double where counties whose coverage is increasing instead decline at the national average and declines double in all counties experiencing declines. (b) Vaccination coverage universally declines 50% more than current trends where the scenario is the same as (a) but instead of declines doubling they are multiplied by 1.5. (c) All counties reach 95% coverage by 2030 by linearly increasing their coverage if currently below 95%. (d) No change in vaccine coverage from 2025 data.

### Supplementary Tables

##### Table S1: Relationship between within-county variability in MMR vaccine coverage and spatially related outbreaks.

| Description | Interaction coefficient | MSR p-value | R² (%) |
|---|---|---|---|
| Base model: SD × Outbreak size | -0.00011 | 0.01 | 9.4 |
| Controlling for the fraction of schools that are public | -0.00011 | 0.01 | 13.0 |
| Controlling for state-fixed effects | -0.000073 | 0.1 | 47.3 |
| Controlling for mean MMR vaccine coverage | -0.00011 | 0.01 | 9.4 |
| Alternative measure of variability: CV instead of SD | -0.000063 | 0.02 | 8.3 |
| Alternative measure of outbreak size: log(outbreak size + 1) instead of outbreak size | -0.0022 | 0.08 | 11.1 |

Coefficients describe the change in the posterior probability of correlation between neighboring counties per unit change in an interaction term of the variability of the within-county MMR vaccine coverage and outbreak size. Negative values indicate that increasing the variability (for a constant outbreak size) decreases the probability of correlation between neighboring counties, consistent with the hypothesis that multi-county outbreaks are associated with homogeneous susceptibility. Each row presents an alternative sensitivity analysis model. The base model is an interaction version over all outbreak sizes of what is presented in the main text. All *p* values are calculated empirically by obtaining a null distribution with equivalent Moran spectral autocorrelation as is between counties in the data.

##### Table S2: Sensitivity analyses of future projections under modified priors.

| Future vaccination scenario | Projected cumulative cases, 2026-2030 [95% Credible interval] |
|---|---|
| Vaccine coverage remains at 2025 levels | 8,000 [2,400, 36,600] |
| Vaccination trends continue | 66,900 [27,900, 158,400] |
| Vaccine declines accelerate 2x | 221,600 [93,000, 510,200] |
| Vaccine declines accelerate 1.5x | 114,700 [46,800, 258,200] |
| All counties reach 95% coverage by 2030 | 2,200 [900, 10,500] |
| 1 percentage point increase in all counties every year | 5,500 [1,700, 21,700] |

We reran the validation and future projection analyses using wider priors (Table S4) and present estimates for future cases using the refit model (best-performing validation model: statistical model with all covariates, state-specific vaccine coefficients, and a conditional autoregressive term at the county-level with training data since 2019). There is considerable overlap in the 95% credible intervals between both the primary model fits and this sensitivity analysis. The model is fit in log space, so small changes to parameter estimates in log space may results in larger changes to projected cases.

##### Table S3: Estimated relationship between nonmedical exemption policies and changes in MMR coverage.

| State | Year | Slope (95% CI) | Intercept | R² (%) | N |
|---|---|---|---|---|---|
| California only | Policy change year: 2016 | 0.745 [0.533, 0.955] | 0.425 | 49.67 | 56 |
| California only | 1 year after: 2017 | 0.683 [0.442, 0.842] | 0.289 | 30.09 | 57 |
| California only | 2 years after: 2018 | 0.665 [0.412, 0.857] | 0.237 | 22.45 | 57 |
| Connecticut only | Policy change year: 2022 | 0.914 [-0.75, 1.667] | -0.316 | 56.29 | 8 |
| Connecticut only | 1 year after: 2023 | 0.5 [-0.636, 1.467] | 0.975 | 41.48 | 8 |
| Maine only | Policy change year: 2021 | 0.362 [-0.266, 1.046] | 0.586 | 17.16 | 16 |
| Maine only | 1 year after: 2022 | 0.628 [0.071, 1.321] | -0.57 | 28.90 | 16 |
| Maine only | 2 years after: 2023 | 0.798 [-0.269, 1.997] | -0.982 | 25.31 | 16 |
| New York only | Policy change year: 2019 | 0.754 [0.535, 0.955] | 0.117 | 31.19 | 61 |
| New York only | 1 year after: 2020 | 1.015 [0.867, 1.144] | 0.049 | 33.92 | 61 |
| New York only | 2 years after: 2021 | 0.914 [0.731, 1.061] | 0.062 | 27.97 | 61 |

| | | | | | |
|---|---|---|---|---|---|
| All states | Policy change year | 0.697 [0.556, 0.850] | 0.275 | 48.47 | 141 |
| All states | 1 year after | 0.725 [0.611, 0.819] | 0.342 | 35.33 | 142 |
| All states | 2 years after | 0.704 [0.582, 0.826] | 0.259 | 25.61 | 134 |
| All states with state fixed effects | Policy change year | 0.713 | 0.452 | 46.57 | 141 |
| All states with state fixed effects | 1 year after | 0.745 | 0.275 | 23.89 | 142 |
| All states with state fixed effects | 2 years after | 0.759 | 0.044 | 16.65 | 134 |

All regression slopes and intercepts are from a Theil-Sen robust estimator that calculates slope and intercepts separately. Uncertainty intervals are the 95% uncertainty range from the Theil-Sen robust estimator.

**Descriptions of Supplementary Data**

**Supplementary Data S1: Model selection metrics for all models on out-of-sample validation performance from 2021-2025.**

**Supplementary Data S2: Out-of-sample classification performance for all models in predicting whether a county will have at least a given number of cases.** (a) Performance predicting whether a county will have at least a given number of cases, including international importations. (b) Performance predicting whether a county will have at least a given number of cases excluding known international importations and therefore validated on counties that observed at least one case.

**Supplementary Data S3: Out-of-sample regression performance metrics for all models in predicting county-level case counts.** (a) Performance predicting total county-level cases, including international importations. (b) Performance predicting cases excluding known international importations and therefore validated on counties that observed at least one case.

**Supplementary Data S4: Model-estimated measles outbreak risk percentiles and international importation probabilities for all counties and states.** (a) County-level table. (b) State-level table.

**Supplementary Data S5: Model-estimated nationwide measles cases under future vaccine and intervention scenarios.**

**Supplementary Data S6: Model-estimated change in outbreak risk from targeted nonmedical exemption policies.** (a) County-level table. (b) State-level table.
